# Trio-based GWAS reveals novel loci associated with different forms of isolated cleft lip

**DOI:** 10.64898/2026.03.15.26348427

**Authors:** Noah Herrick, Zeynep Erdogan-Yildirim, Myoung Keun Lee, Sarah W Curtis, Seth Berke, Grace Brewer, Toby McHenry, Ahmed M El Sergani, Joel Anderton, Nandita Mukhopadhyay, Jenna C Carlson, Terri Beaty, Azeez Butali, Carmen J Buxo-Martinez, Jacqueline T Hecht, Eric Liao, Lina M Moreno Uribe, Carmencita D Padilla, George Wehby, Eleanor Feingold, Jeffrey C Murray, Ingo Ruczinski, Elizabeth J Leslie-Clarkson, Seth M Weinberg, John R Shaffer, Mary L Marazita

**Author notes:** Corresponding author: Noah Herrick, PhD, Center for Craniofacial and Dental Genetics, Bridgeside Point 1, Suite 400, 100 Technology Drive, Pittsburgh, PA 15219,; Mary L. Marazita, PhD, Center for Craniofacial and Dental Genetics, Bridgeside Point 1, Suite 400, 100 Technology Drive, Pittsburgh, PA 15219. Shared first author (Noah Herrick, Zeynep Erdogan-Yildirim).

## Abstract

Orofacial clefts (OFCs) are the most common craniofacial birth defect and comprise a diverse group of traits with complex and heterogeneous etiologies. Genetic studies of OFCs typically approach this diversity by stratifying cases into broad diagnostic classes, including cleft lip (CL), cleft palate (CP), and cleft lip with palate (CLP). Although this strategy has yielded important insights into OFC risk, it ignores the phenotypic heterogeneity within each subtype. CL exhibits marked phenotypic variability, involving differences in alveolar involvement, laterality, and sidedness that may reflect distinct etiologies. Given this phenotypic diversity within CL, we assembled a multi-ancestry cohort of 837 nonsyndromic CL case-parent trios with whole-genome sequencing and detailed phenotyping. We performed genome-wide association scans (GWAS) via transmission disequilibrium tests for CL overall and for 14 CL subtypes defined by involvement of the alveolus (with and without), laterality (uni- and bilateral), and sidedness (left and right). We identified four genome-wide significant loci. Two loci, *IRF6* and 8q24.21, were both detected in the overall CL GWAS. *PLCB1*/*PLCB4* and *MAFB* were detected in GWASs of alveolar cleft involvement and CL left sidedness, respectively. These subtype-specific associations were followed by case-only comparisons that reflect the presence or absence of alveolus cleft or left-sided bias of CL to confirm the specificity of the association signal to the particular subtype. Our results provide additional, new evidence of CL subtype-specific genetic links for loci previously discussed in the context of primary OFC classes and demonstrate the value of granular OFC subtype characterization to capture trait-specific associations.

## INTRODUCTION

Orofacial clefts (OFCs) are among the most prevalent congenital malformations, affecting approximately 1 in 700 live births worldwide^1^. Isolated OFCs, also referred to as nonsyndromic OFCs, are not accompanied by other structural or developmental anomalies and are far more common, accounting for roughly 70% of all OFC cases^1^. Unlike syndromic forms, isolated OFCs have a complex and multifactorial etiology^2,3^. Twin studies show evidence of high heritability, consistent with elevated familial recurrence risk^4–6^ and genome-wide association studies (GWAS) have identified over 50 risk loci to date^7–18^. Combined, these loci only partly explain the estimated heritability, leaving a gap in our knowledge of the genetic basis of isolated OFCs.

Isolated OFCs can show a wide range of phenotypic presentations, varying by location and severity. There is also emerging evidence that distinct patterns of genetic association may underlie different forms of isolated OFC. As a consequence, the use of accurate and detailed phenotypes in genetic studies of isolated OFCs is critical. For affected individuals, OFCs have been mainly classified into cleft lip (CL), cleft palate (CP), and the combined condition of cleft lip with cleft palate (CLP). CL and CLP are often treated as a single entity in most genetic studies (termed as CL/P), with CLP regarded as the more severe form^8,9,12,19–26^. However, recent GWASs have shown that CL and CLP each demonstrate distinct genetic risk factors^16–18,20,27–32^. In a recent study, our group further refined the proband’s subtype definition (e.g. CL) by incorporating the cleft types observed in their extended family (CL or CLP or mixed), yielding more genetically homogeneous subsets and improved discovery of subtype-specific genetic associations despite reduction in the sample size^33^.

Notably, CL itself is a quite heterogenous phenotype featuring variation in alveolar bone involvement, laterality (unilateral versus bilateral), and sidedness (right versus left). Despite growing evidence that the major isolated OFC subtypes exhibit distinct genetic risk patterns^20,34–36^, the genetics of isolated CL and its various phenotypic presentations remain significantly understudied—largely due to limited high-resolution phenotyping and small subgroup sample sizes with available genotypic data. Although multiple GWASs have investigated CL^11,17,18,27,33^, fewer studies have incorporated the trait’s full range of phenotypic features—such as alveolar involvement^34,35^, laterality^30^, and sidedness^20^. To address this critical research gap, in this study we leverage whole genome sequencing (WGS) data from the largest cohort of CL trios to date which includes 837 trios comprising multiple ancestral backgrounds with comprehensive phenotyping available through the Gabriella Miller Kids First (GMKF) Pediatric Research Program.

## METHODS

### Discovery study population

Our isolated CL study sample combined existing WGS data from case-parent trios obtained from six separate GMKF cohorts (n=837). The details of these cohorts, including ancestral background distributions, are provided in Tables S1 and S2. All participants in this study provided individual informed consent under recruitment and study protocols approved by their local Institutional Review Boards (IRBs) or Ethical Committees and overseen by the study Coordinating Center at the University of Pittsburgh. More details and Federal Wide Assurance (FWA; USA accreditation of IRBs) numbers are provided in the Ethical Oversight section.

### CL Phenotyping

Trios were only included if the case presented with isolated CL. Family history of other cleft types (e.g., a relative with CLP) did not impact inclusion criteria for the proband with CL. The LAHSHAL system was used to collect detailed information about the cleft status of affected cases^37,38^ and their family members at each of the recruitment sites by trained, calibrated, and experienced field staff. Information about alveolar involvement (lip only or lip and alveolus), laterality (unilateral or bilateral presentation), and sidedness for unilateral clefts (left or right) was noted. The availability of these detailed phenotypic data (dbGaP) allowed us to partition cases (and trios) into 15 different but not mutually exclusive CL subtypes (Figure 1), based on alveolar involvement, cleft laterality (unilateral or bilateral), and sidedness (left or right sided).

**Figure 1.**
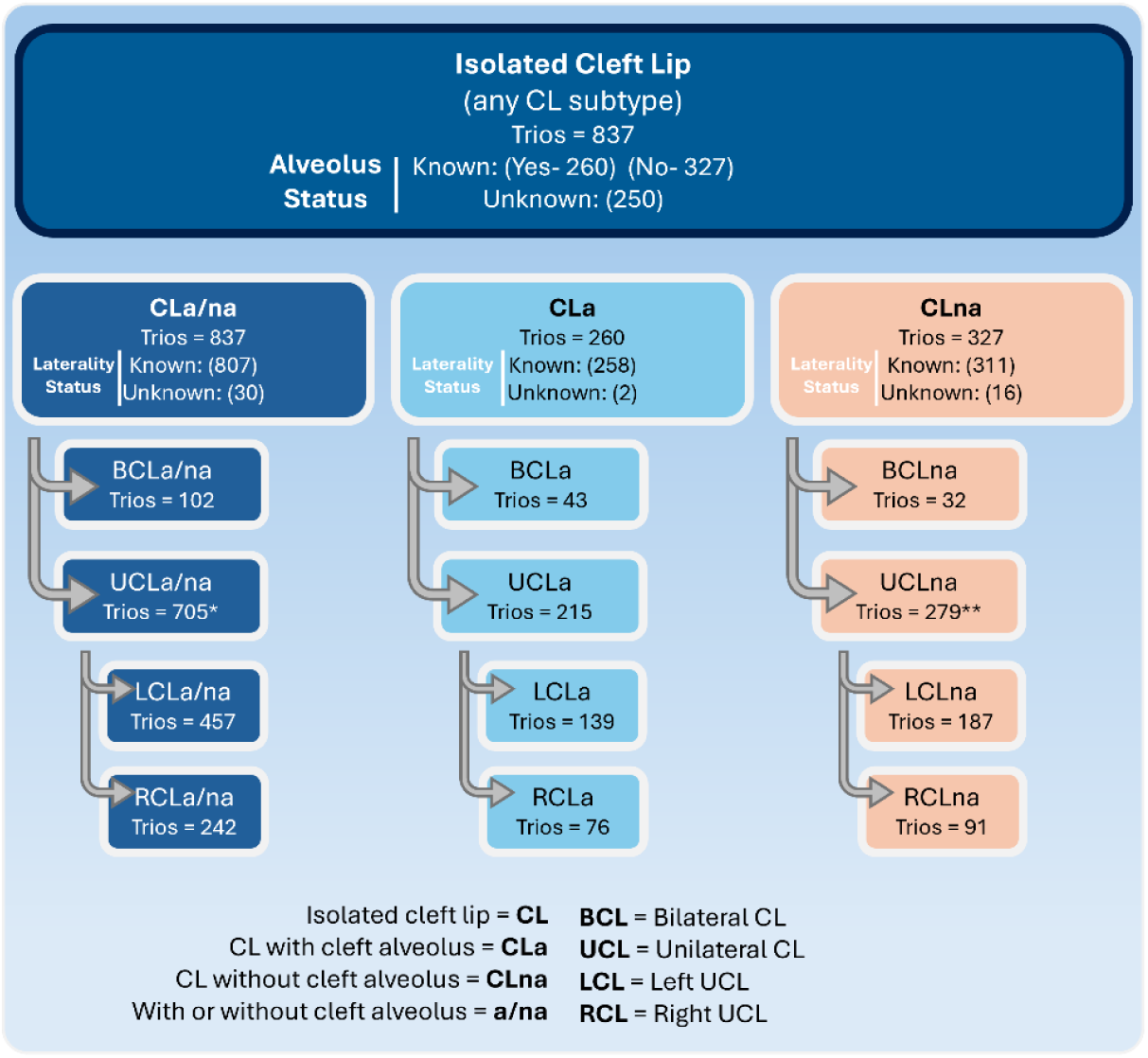
Diagram of CL case-parent trio classification into appropriate subtypes. CLa/na includes all possible CL subtypes in this analysis, independent of alveolus or laterality status. Probands with ambiguous CL subtypes are also included in this all-inclusive set. Probands that have a known status for cleft alveolus (yes/no) are subdivided into CLa (yes) and CLna (no). All sample sizes provided represent the number of case-parent trios. *There are six trios included in UCLa/na with unknown sidedness status. **There is one trio included in UCLa/na with unknown sidedness status.

The broadest category was isolated CL with or without alveolar involvement (CLa/na; 837 trios) which included all subtypes. Then, we divided this based on alveolar involvement to CLa (CL with cleft alveolus; 260 trios) and CLna (CL without cleft alveolus 327 trios). Within each of these three phenotype definitions we further subdivided into bilateral (BCL) and unilateral (UCL). Lastly, we also subdivided UCL based on sidedness into right unilateral CL (RCL) and left unilateral CL (LCL). Illustrations of phenotypic manifestation of these CL subtypes are shown in Figure 2.

**Figure 2.**
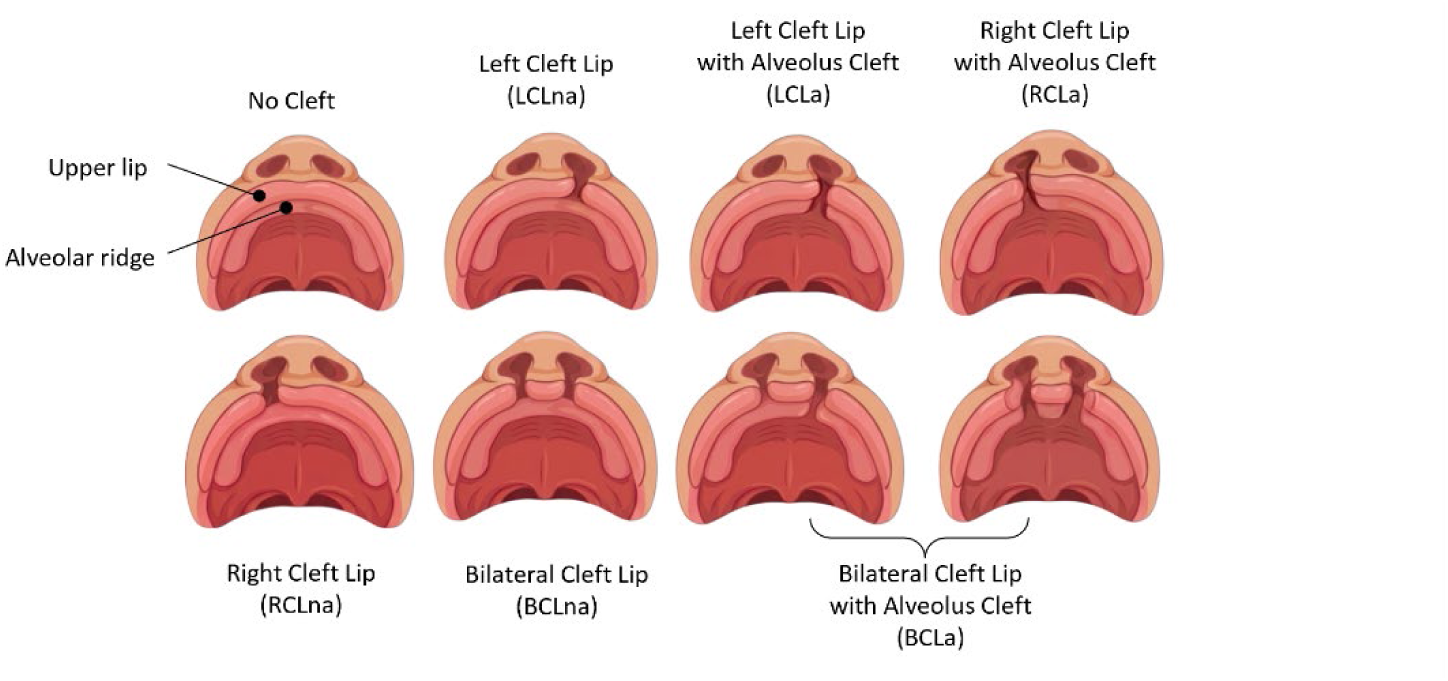
While OFCs are a heterogenous collection of CL, CP, CLP, and CL/P, CL is itself heterogenous and can present in various CL subtypes. Note, the BCLa subtype included probands with alveolar involvement regardless of alveolus-specific laterality (including uni- or bilateral alveolus cleft) or alveolus-specific sidedness (left-sided alveolus cleft is shown above; right-sided alveolus cleft is not shown above but probands were included). Images were generated with BioRender.

### Short-read sequencing and quality control

DNA samples from study participants were subjected to short-read whole-genome sequencing by either the McDonnell Genome Institute at Washington University School of Medicine in St. Louis or the Broad Institute, all targeted to achieve an average of 30X coverage. Variant calling on GMKF trios were aligned to the GRCh38 genome reference assembly, using harmonization techniques in recommended GATK pipelines^39–41^ (https://software.broadinstitute.org/gatk/best-practices/workflow). The GMKF pipelines are open source and made available to the public via GitHub (alignment workflow: https://github.com/kids-first/kf-alignment-workflow; joint-genotyping workflow: https://github.com/kids-first/kf-jointgenotyping-workflow).

We also applied quality control metrics with BCFtools (v1.22) to decompose complex variants and split multi-allelic sites into biallelic records (norm -a -m -any). We filtered out genotypes with either a Phred-scaled probability (QUAL) scores less than 20 or a read depth (DP) less than 10 in tandem with a genotype quality (GQ) less than 20 (filter -e QUAL<20 || FMT/DP<10 & FMT/GQ<20’). After these steps were performed on each dataset, all six datasets were merged (-merge) and subjected to more quality control prior to trio-based mega-GWASs, mentioned hereafter. The mega-GWAS approach was best suited for combining the several smaller trio cohorts with the larger ones. Using PLINK^42^ (v1.90), all SNPs with a missing rate greater than 5% or a Mendelian error count greater than one were removed. Individuals that presented Mendelian error rates greater than 2% on any given chromosome prompted exclusion of the corresponding case-parent trio set (18 trios were dropped, resulting in 837 trios). A minor allele frequency threshold greater than or equal to 3% was used to keep common variants. Hardy-Weinberg equilibrium (HWE) was assessed separately for the three largest populations (based on self-report) in our dataset. These populations corresponded with distinct genetic principal component (PC) clusters representing African, Asian, and European ancestry seen in PC1 (Figure S1). Markers were excluded if HWE test P-values were less than or equal to the 1e-6 threshold in at least one of the three populations. These quality control measures provided a final set of 6,065,803 common genetic variants for analysis of CLa/na and similar marker sets in the other subtypes (Table S3).

### Statistical analyses

Transmission disequilibrium tests (TDTs) were performed with PLINK^42^ (v1.90) for each of the 15 CL subtypes to evaluate evidence for genetic association. We report associations based on genome-wide significance (P < 5e-8) and suggestive (P < 5e-6) thresholds, as well as a study-wide significance threshold (P < 3.3e-9) calculated using a conservative Bonferroni correction (5e-8 / 15 phenotypes). A detailed summary of these phenotypic subgroups and sample sizes for each of these corresponding tests is provided in Figure 1. The TDT is robust to population stratification and does not require statistical adjustment for ancestry. However, we did perform genetic PC analysis using PLINK (v1.90) to observe the population structure of our isolated CL cohort compared to self-reported ancestral background (Figure S1), as well as adjust for population structure in the follow-up case-case logistic regression analyses.

Next, we performed genetic modifier tests (case versus case logistic regressions via PLINK^42^) on lead SNPs with possible subtype specificity to validate the observed genetic associations with specific CL subtypes. This test can detect genetic risk factors that vary between two groups and has shown prior success with OFC subtypes^20,30,31^. The first four genetic PCs (totaling more than 98% of variance) were used as covariates in this SNP candidate genetic modifier approach to adjust for population structure. There were known CL sibling probands in case-parent trios, therefore only one CL case was randomly selected for the case-only genetic modifier tests totaling 788 total unrelated CL cases. We also used KING-robust kinship estimator^43^ to ensure there were no unknown relationships. The sample size for each case-only subgroup is detailed in Table S4.

### Gene annotation and fine mapping

The regional genomic associations (1 MB window) for the independent loci were visualized with GWASLab^44^ and an Ensembl gene track (GRCh38). Lead SNPs were mapped to genes using the three gene mapping strategies based on genomic position, eQTL data, and chromatin interactions implemented in FUMA^45^ (v1.5.2) and detailed in File S1. The isolated CL subtype where the lowest P-value was observed for the lead SNP was the input for FUMA. Additional configurations and parameters for the FUMA analyses are also provided in the supplement (File S1).

To fine-map association signals and prioritize potential causal variants, we applied a Bayesian sparse regression framework using SuSiE, implemented in the susieR package^46^ (v0.14.2) and integrated through GWASLab^44^. Fine-mapping was conducted on GWAS summary statistics, incorporating effect size estimates, standard errors, and pan-ancestry linkage disequilibrium (LD) reference panels from the 1000 Genomes Project^47^. Analyses were performed using the ‘susie_rss’ function, permitting up to 10 causal variants per locus (L = 10; default). The credible set (CS) threshold was set at 95% coverage. The default prior variance parameters were used, and all analyses were restricted to ±50 kb windows centered on the lead SNP for each locus.

### Replication of known Isolated cleft lip signals

CL-significant SNPs from 27 loci previously reported in literature^11,17,18,27,33^ were selected for replication tests in the all-inclusive CLa/na TDT. We converted any hg19 genomic locations of those SNPs from literature to hg38 with liftOver (v1.24.0) for equivalent comparison to our work. If there were exact SNP matches not available in our dataset, which occurred for five loci, we checked proxy SNPs within a ±250kb window and linkage disequilibrium (LD) r^2^ > 0.80 using LDlink, however, those possible proxy SNPs were also absent. A Bonferroni-adjusted threshold (alpha/number of independent loci tested: 0.05/22) was used to determine if the known SNP was replicated in our study.

## RESULTS

We performed 15 GWASs of isolated CL and subtypes (Figures S2-S6). We use the following acronyms to label five phenotypes based on laterality and sidedness: CL = cleft lip (including all laterality and sidedness subtypes); BCL = Bilateral CL; UCL = Unilateral CL (including both left and right sidedness); LCL = Left UCL; and RCL = Right UCL. For each of these five laterality and sidedness phenotypes we use three suffixes to represent alveolar involvement: “a” = with cleft alveolus, “na” = without cleft alveolus, and “a/na” = with or without cleft alveolus which includes those with an unknown status.

### GWAS of isolated cleft lip (CLa/na)

There were two significant loci with lead SNPs surpassing the genome-wide significance threshold (P < 5e-8; Figure 3a; Figure S7). Each locus, *IRF6* and 8q24.21, has previously been associated with isolated CL and isolated CLP. There were 16 suggestive loci (P < 5e-6; Table S5). We attempted to replicate 22 previously associated CL loci from multiple studies^11,17,18,27,33^, and six (27%) surpassed Bonferroni-adjusted significance (P < 0.05/22) in this study (Table S6).

**Figure 3.**
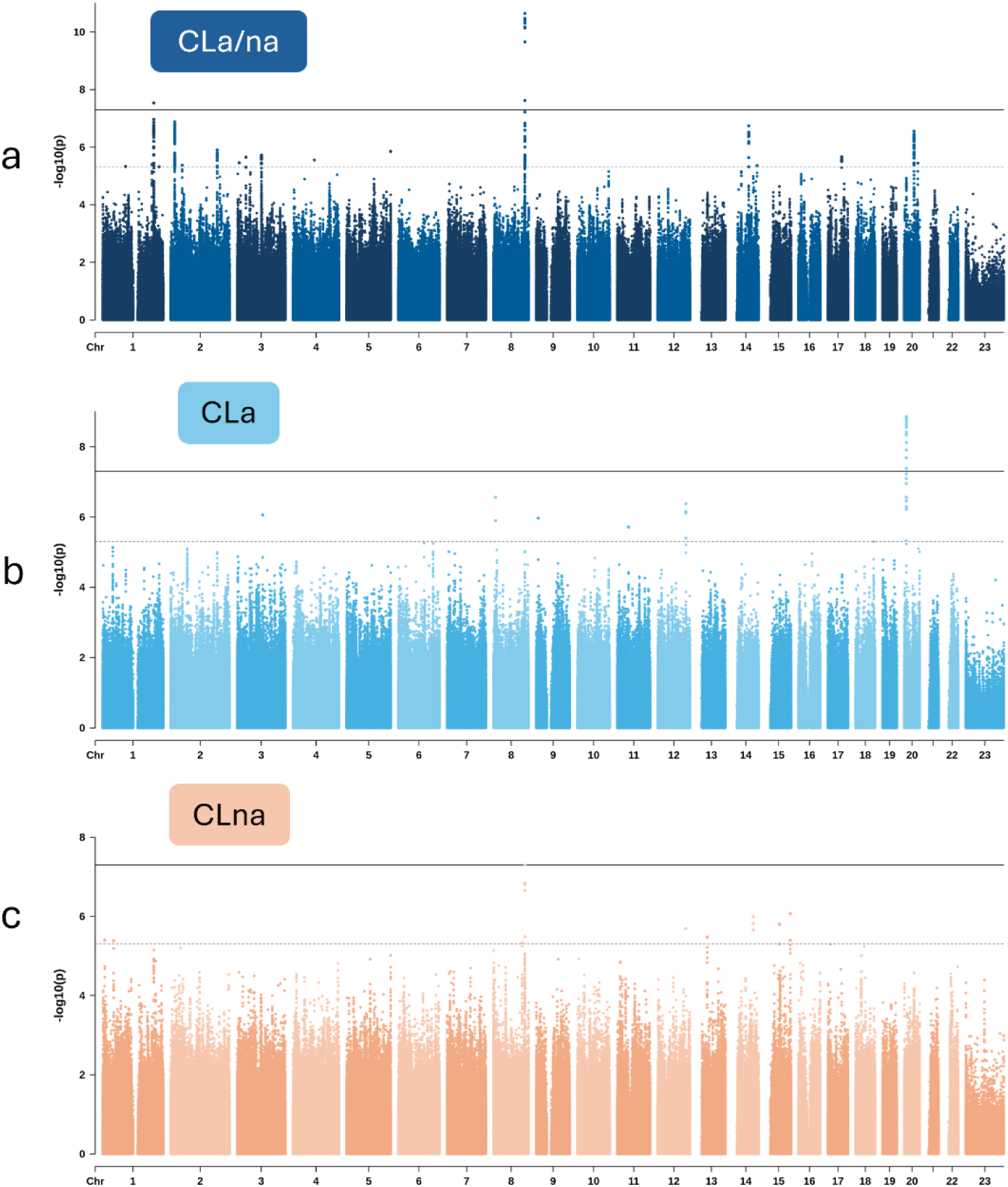
Manhattan plots of the GWASs for the three primary groupings of CL in this study: **a)** CLa/na includes all CL subtypes and unknown statuses of CL subtypes (trios=837), **b)** CLa includes trio sets with probands presenting with known cleft alveolus and independent of laterality (trios=260), and **c)** CLna includes trio sets with probands presenting with known absence of cleft alveolus (trios=327).

### GWAS of cleft lip subtypes

Two loci displayed genome-wide significant associations with CL subtypes. One locus, located in an intron of *PLCB1* upstream of *PLCB4*, highlights a genetic association observed for CLa (Figure 3b,c). This association is present in the largest CLa grouping which includes all laterality and sidedness subtypes (P = 1.40e-9) but was strongest in UCLa (P = 1.08e-9). Each of these two subtype associations with *PLCB1*/*PLCB4*, CLa (n = 260) and UCLa (n = 215), achieved study-wide significance. Within the strongest associated subtype, UCLa, lead SNP rs2206265 presented consistent allelic transmission rates across ancestry groups (Table S7). The same locus showed an absence of association in all CLna subtypes. The lead SNP’s P-value in CLna (n = 327), for example, did not reach nominal significance (P = 0.61). Regional association plots were used to illustrate the contrast of this association between CLa/na, CLa, and CLna (Figure 4).

**Figure 4.**
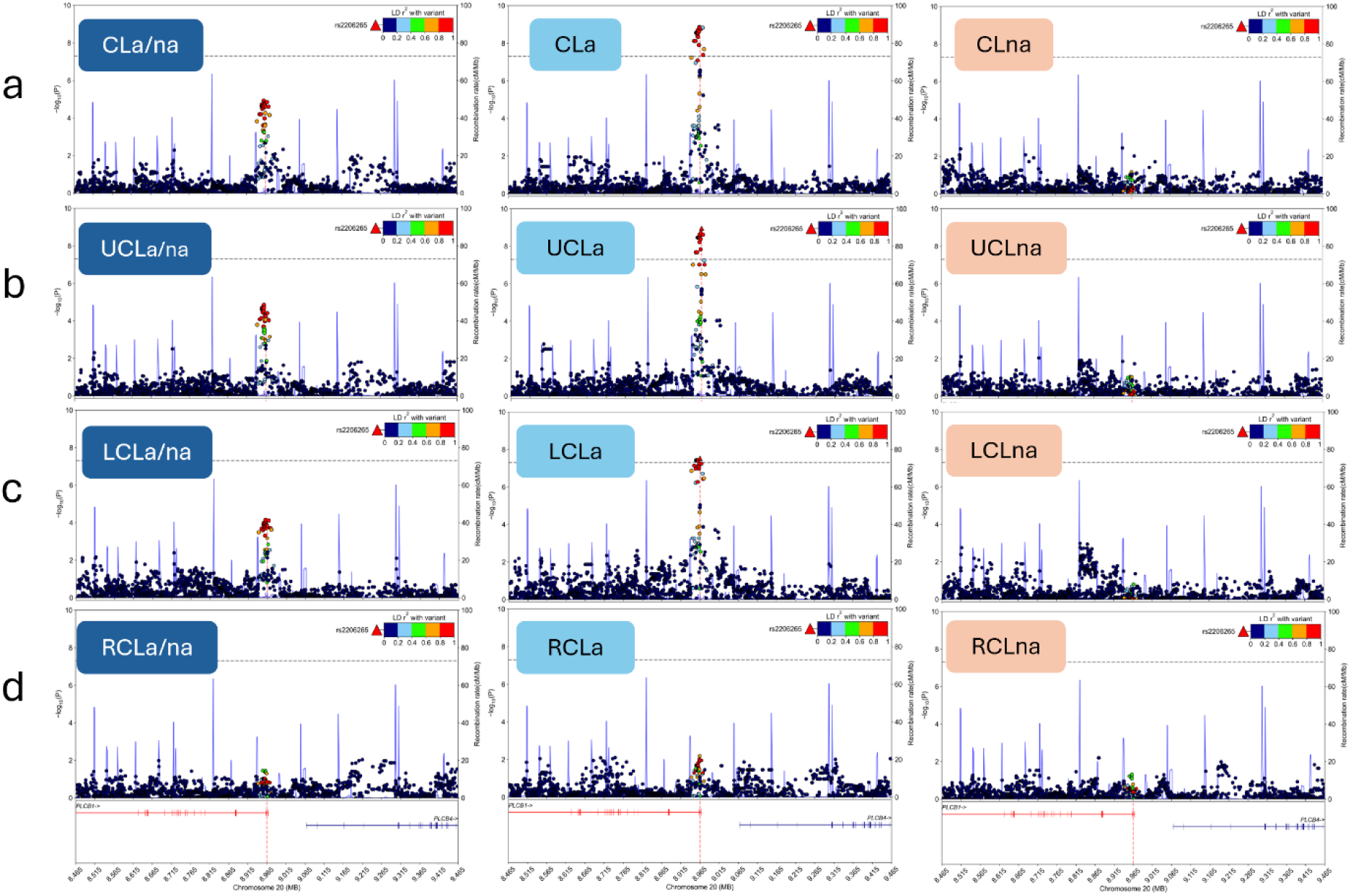
Regional association plots of the alveolus cleft subtype-specific signal, *PLCB1*/*PLCB4*. The column order for these plots is based on alveolar involvement: all-inclusive (a/na), alveolus cleft present (a), and alveolus cleft absent (na). The row order is based on laterality and sidedness: **a)** all-inclusive (CL), **b)** unilateral, **c)** left unilateral, and **d)** right unilateral. The bilateral subtypes were excluded from these plots on the basis of very small sample sizes.

The summary statistics from the GWAS of UCLa were used as input for FUMA (v1.5.2) to perform three gene mapping approaches based on genomic location, eQTL data, and chromatin interactions. All three strategies were concordant in mapping this locus to PLCB4 (Figure S8c).

Available Hi-C data on a UCSC Genome Browser craniofacial track hub indicated that both genes, PLCB1 and PLCB4, are located in the same cranial neural crest cell (CNCC) topological associated domain (TAD) which also includes a craniofacial super-enhancer region (Figure S9) that is predicted to interact with the promoters of both genes. Two of the significant variants from this locus (non-lead SNPs) are found in the same candidate cis-regulatory element (cCRE) classified as a transcriptional start site (TSS) distal enhancer. Fine-mapping tool, SuSieR (v0.14.2), identified a single causal signal in this locus with 19 of the genome-wide significant SNPs (76%) to be in the 95% credible set, with the lead SNP, rs2206265, highlighted with the highest posterior inclusion probability (PIP = 0.13).

The second genome-wide signal, comprising intergenic variants upstream of *MAFB*, was associated with LCLa/na (P = 8.41e-9). This lead SNP, rs6065259, also presented consistent allelic transmission rates across ancestry groups (Table S7). While this same locus also appeared as a suggestive signal in the UCLa/na TDT (P = 1.26e-7), there was no signal observed from the RCLa/na GWAS (Figure 5). This contrast in sidedness association, left versus right, is shown clearly in Figure 6, where the lead SNP showed no association with RCLa/na (P = 0.28). The same FUMA-based gene mapping approach as above demonstrated eQTL connections with *EMILIN3* and several chromatin interactions including those with the nearest gene, *MAFB* (Figure S8d). The genome-wide significant SNPs in this locus (n=3) are positioned in the same CNCC TAD, two of which overlap with cCREs: either a distal enhancer (lead SNP, rs6065259) or an element with high chromatin accessibility (CA) and CCCTC-binding factor (CTCF) activity (CA-CTCF; rs6072082; Figure S10). Fine-mapping tool, SuSieR (v0.14.2), identified a single causal signal in this locus with all three of the genome-wide significant SNPs (100%), in addition to 22 other SNPs in high LD, to be in the 95% credible set. The lead SNP, rs6065259, resulted in the highest PIP (0.24) of the credible set.

**Figure 5.**
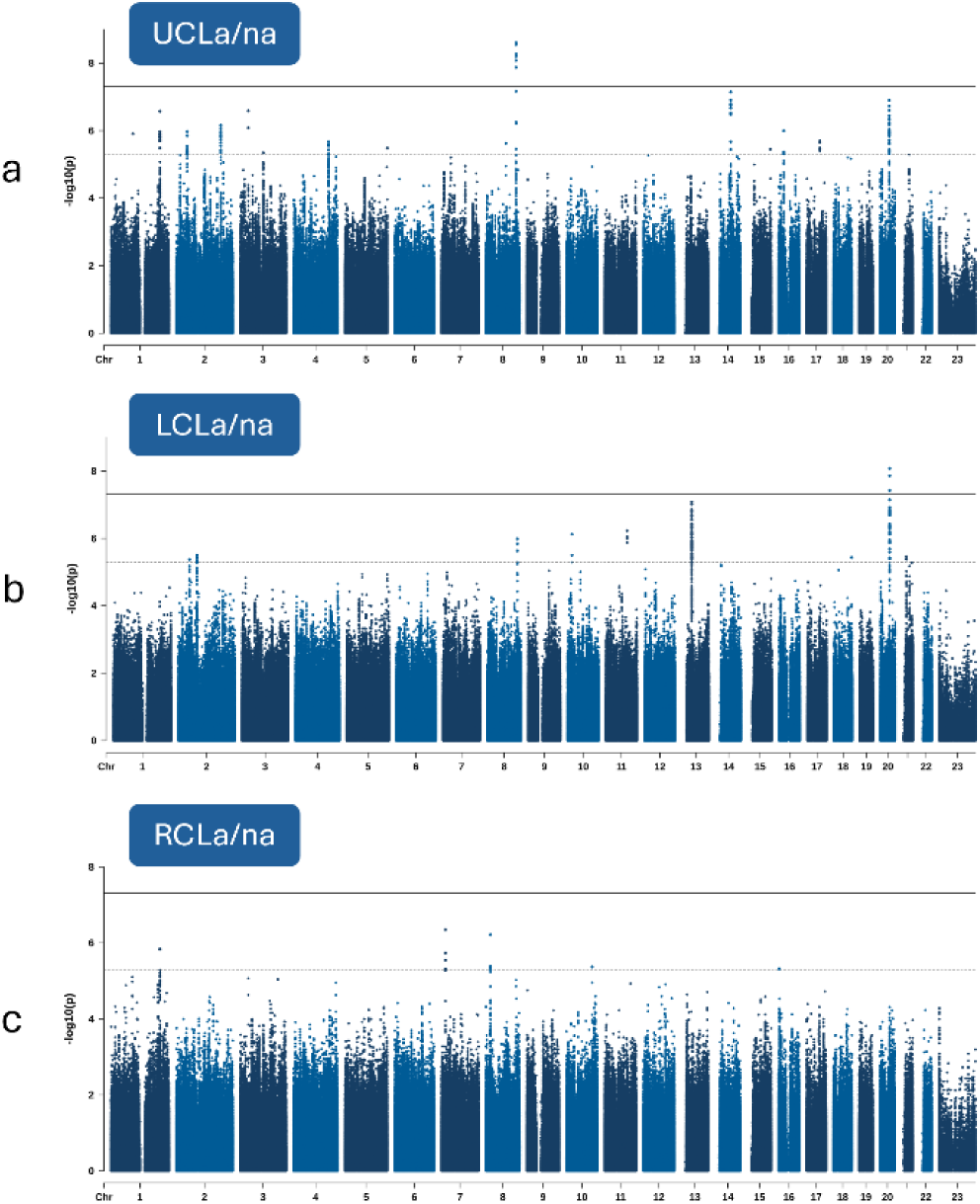
Manhattan plots of the GWASs for three subtypes of CLa/na in this study based on sidedness: **a)** UCLa/na includes unknown sidedness statuses (trios=6) of UCL subtypes (trios=705), **b)** LCLa/na includes trio sets with probands presenting with known left-side CL and independent of alveolus cleft status (trios=457), and **c)** RCLa/na includes trio sets with probands presenting with known right-side CL and independent of alveolus cleft status (trios=242).

**Figure 6.**
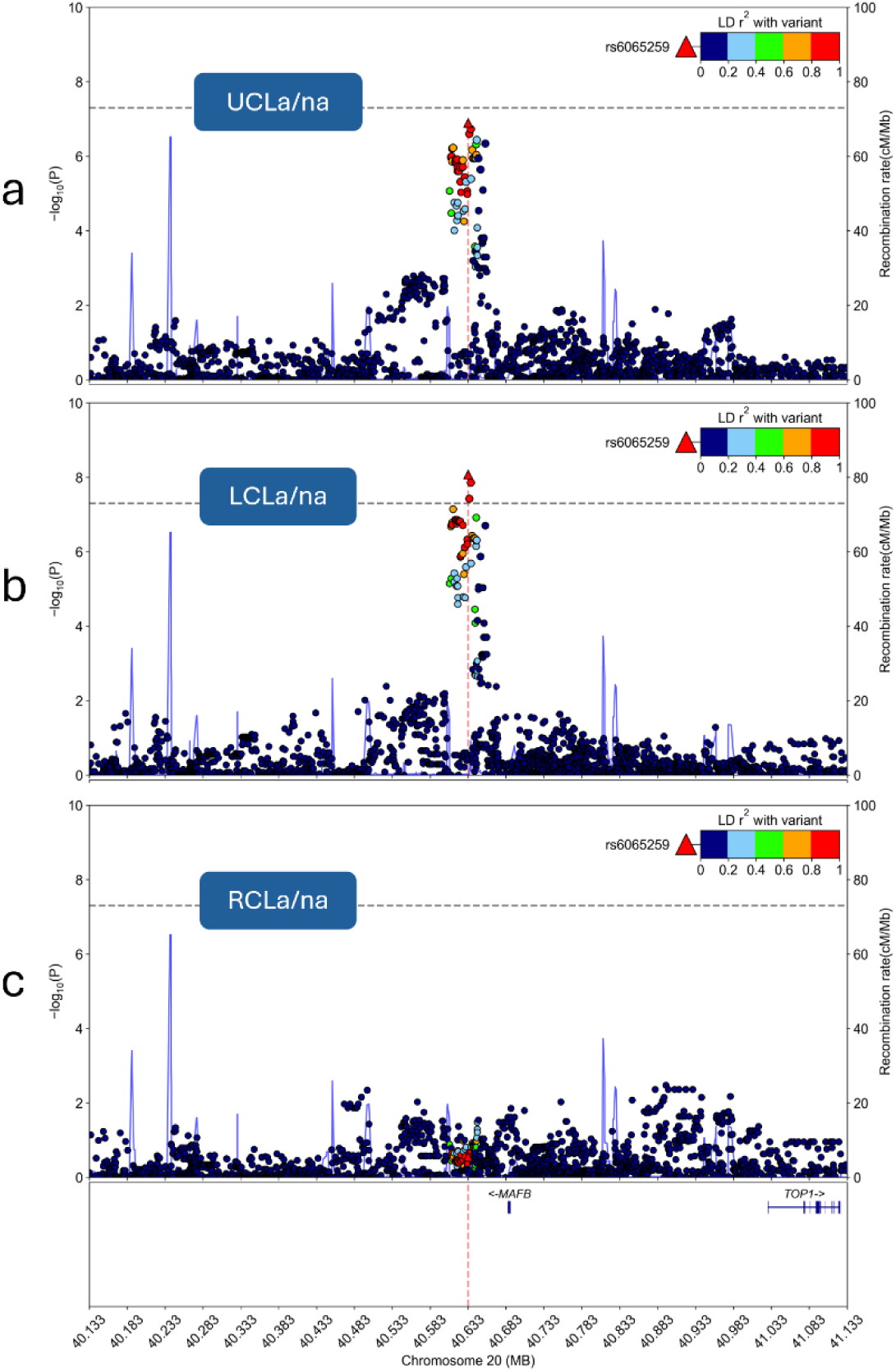
Regional association plots of the sidedness cleft subtype-specific signal near MAFB for **a)** all UCLa/na subtypes and unknown sidedness statuses of UCL subtypes, **b)** left-side CL with or without alveolus cleft, and **c)** right-side CL with or without alveolus cleft.

### Effect estimates and modifier analysis of gene candidates

We generated forest plots of odds ratios (ORs) and 95% confidence intervals (CIs) for each of the four lead SNPs presented in this study to visualize the subtype-specificity of the associations (Figure 7a). The ORs for both lead SNPs in the CLa/na GWAS, rs72741048 and rs17242358, are not subtype-specific, and the effect alleles for each SNP show consistency in direction of effect across all subsets of CLa/na. Effect estimates for the third lead SNP, rs2206265, presents strong evidence for alveolus cleft specificity with high ORs only for subtypes involving the alveolus. In contrast, all subtypes without alveolar involvement showed ORs closer to one. Likewise, the fourth SNP, rs6065259, sheds light on unilateral cleft sidedness favoring the left versus the right. The ORs for left sided subtypes (LCLa/na, LCLa, and LCLna) are approximately 0.5 showing a strong protective effect, whereas the ORs for the right sided subtypes (RCLa/na, RCLa, and RCLna) are close to one.

**Figure 7.**
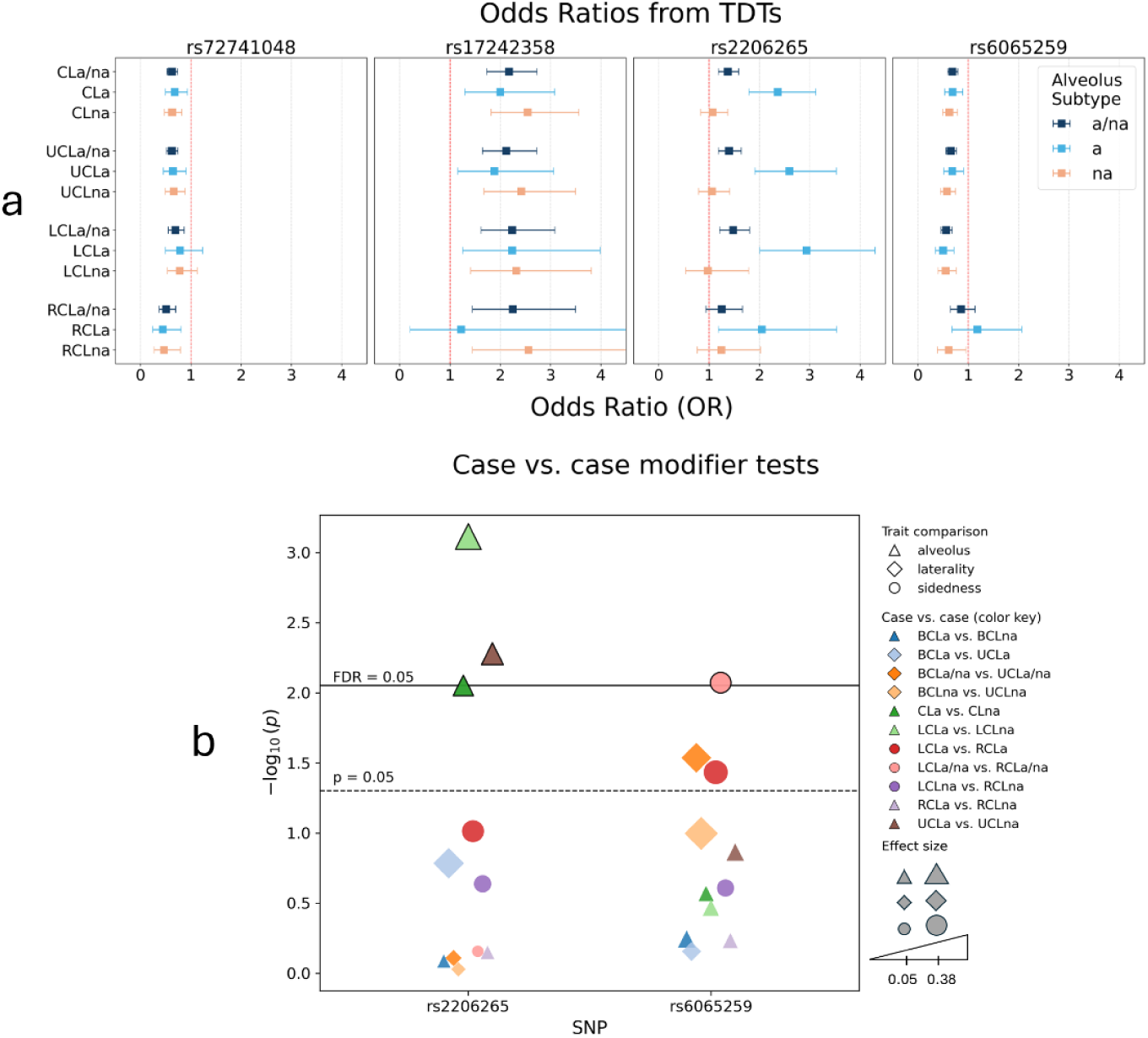
Effect estimates and case versus case modifier analyses of lead SNPs with **a)** ORs and 95% CIs from the original TDTs for all four lead SNPs across all subtype-specific analyses excluding bilateral subtypes on the basis of very small sample sizes and consequently unreliable ORs and CIs. The ORs are color coded by alveolus cleft subtype (a/na: dark blue), (a: light blue), and (na: light orange). **b)** Genetic modifier tests for the two lead SNPs with subtype-specific associations. Shapes represent the subtype-specific trait being compared (triangle: alveolus cleft; diamond: laterality; circle: sidedness). The colors illustrate which exact comparison was performed. The size of the shape represents the effect size resulting from the case versus case test. Thresholds represent nominal significance (P = 0.05) and FDR-adjusted significance (FDR = 0.05).

To further examine the two lead SNP candidates showing evidence of subtype specificity, rs2206265 (*PLCB1*/*PLCB4* for alveolar involvement) and rs6065259 (*MAFB* for sidedness), we performed case versus case association tests contrasting 11 pairs of subtypes (Figure 7b). These comparisons are sensitive to identifying variants that may be important for one subtype compared to the other. We selected specific pairwise comparisons that either tested alveolar involvement (e.g., LCLa versus LCLna), laterality (e.g., BCLa/na versus UCLa/na), or sidedness (e.g., LCLa/na versus RCLa/na). Note, case versus case tests can only be performed for mutually exclusive groups. Lead SNP rs2206265 showed FDR-adjusted significant differences in LCLa versus LCLna (P = 7.69e-4; FDR = 1.69e-2), UCLa versus UCLna (P = 5.27e-3; FDR = 4.88e-2), and CLa versus CLna (P = 8.87e-3; FDR = 4.88e-2), indicating its specificity in alveolus cleft formation. The other lead SNP, rs6065259, showed an FDR-adjusted significant difference in LCLa/na versus RCLa/na (P = 8.48e-3; FDR = 4.88e-2) and nominally significant differences in BCLa/na versus UCLa/na (P = 2.91e-2; FDR =0.13) and LCLa versus RCLa (P = 3.69e-2; FDR = 0.14), adding evidence for the specificity of this variant with left-side CL.

## DISCUSSION

Isolated OFCs are widely recognized as a heterogeneous set of congenital facial anomalies. To address this diversity, studies on isolated OFCs have traditionally stratified analyses by broad diagnostic categories. While this framework has been instrumental in advancing our understanding of OFC risk, it implicitly treats the traditional subtypes as etiologically homogeneous. However, substantial phenotypic variation exists within each of the main subtypes (CL, CLP and CP). Here we focus on CL, where the phenotype can include differences in alveolar involvement, laterality patterns, and sidedness. These features introduce additional layers of etiological heterogeneity and require a more granular approach to reflect possibly distinct underlying genetic mechanisms. To this end, we compiled multiple cohorts of case-parent nonsyndromic CL trios with short-read WGS data and deep phenotypic data from multiple ancestral backgrounds (837 trios) to perform TDTs in the CL group as a whole and in 14 stratified CL subtypes.

The stratified trio-based GWASs provided 39 genome-wide significant SNPs across four independent genomic loci. Despite the size of our all-inclusive CL (CLa/na) analysis, we did not identify any new genome-wide significant signals in this primary group. It is possible that grouping all CL subtypes together may mask some subtype specific associations, even in a larger sample. It is not surprising, though, to have identified two loci, *IRF6* and 8q24.21, well-known for their association to CL^17,18,27^ and observed in previous SNP array GWAS^18^ with a subset of the participants in this study. These two loci were detected at genome-wide significance in the CLa/na GWAS further supporting their link to CL risk. In the UCLa/na GWAS, both the *IRF6* (suggestive) and 8q24.21 (significant) loci appeared as signals. The 8q24.21 loci was also a suggestive association in the CLna GWAS. Both loci showed weak association in the comparable subtype groups with alveolus cleft, CLa and UCLa. Especially with the majority of cases in the ‘a/na’ subgroups presenting without alveolus cleft or having an unknown status, these results paired with evidence in the literature suggest preferential association to CLna in general but do not preclude association to broader categories of OFCs, such as CLP. Our lead SNPs for *IRF6* and 8q24.21 in the CLa/na test were also significant variants in the CL case-control GWAS by Dack et al. ^17^ (2025).

The main novelty of this study emerges as subtype specific genetic associations for alveolar involvement and left-sidedness. Of course, we only investigated isolated CL in this study and future work must explore other classes of OFCs, such as CLP, to consider the presence or absence of these subtype specific associations beyond isolated CL. It is also important to note that these alveolus cleft or left-sidedness signals do not appear to be driven by any single-largest ancestry group as we observe comparable sample sizes, minor allele frequencies, and allele transmission rates across at least two population groups. Nevertheless, the signal we detected in an intronic region of *PLCB1* upstream of *PLCB4* appeared in three of the five GWASs of cases with alveolus cleft. The remaining two GWASs were comparatively underpowered, BCLa and RCLa. Indeed, we must consider the ‘nesting’ of samples in the primary subgroups, where CLa is a combination of BCLa, UCLa, LCLa, and RCLa. Furthermore, UCLa contains all LCLa and RCLa case-parent trios. This could mean the *PLCB1*/*PLCB4* signals in the larger subgroups are being driven by the association to LCLa, for example.

Although our lead SNP is located in an intron of *PLCB1,* and we reflect that in our nomenclature for this study, its genomic positioning in a cCRE distal enhancer and proximity to *PLCB4* along with evidence of eQTL data and chromatin interactions with *PLCB4* raises the possibility of regulatory effects on a well-established craniofacial development signaling pathway. *PLCB4* variants underlying Auriculocondylar Syndrome 2 (ACS2) impair endothelin receptor signaling in CNCCs, producing jaw and oral cavity defects in both human and experimental models^48^. It has also been shown that dominant negative missense mutations in *PLCB4* result in Auriculocondylar Syndrome through disruption of EDN1–DLX5/6–mediated mandibular patterning^49^. ACS2 phenotypes include a variety of mandibular phenotypes including micrognathia, cleft palate and ear abnormalities. Some individuals also have abnormal tissue expansion along the medial alveolar surface such that the mandible takes on a maxillary phenotype, suggestibly through homeotic transformation^49^. Together, these findings suggest that subtle perturbations of *PLCB4*-associated regulatory architecture may influence alveolar cleft phenotypes outside the context of syndromic facial anomalies.

Moreover, *PLCB4* acts on osteoclast-mediated bone resorption and remodeling and osteoblast differentiation^50,51^, which are also important for alveolar bone remodeling^52^. Hence, in the context of alveolar clefting, altered expression of *PLCB4* could impair the intramembraneous ossification at the premaxillary-maxillary suture^53^ and hinder the complete fusion leaving a gap in the alveolus. Since variants in the regulatory region of *PLCB4* were exclusively associated with alveolar clefts in this study, this adds to the growing evidence of a regulatory role of *PLCB4* on alveolar bone fusion. This also warrants further investigation into its possible role in CLP.

The second signal we detected was in an intergenic region upstream of *MAFB*, a transcription factor gene, consistently replicated across OFC GWASs^9,11,54,55^, but phenotypic specificity has been more challenging to pinpoint. Recent functional research demonstrated that neither the cleft-associated *MAFB* H131Q missense variant, rs121912307^56^, nor complete loss of *Mafb* disrupts palatogenesis *in vivo*, indicating that *MAFB* is not required for secondary palate formation^57^. Importantly, this finding does not preclude a role for *MAFB* in CL, which arises from defects in frontonasal and maxillary prominence growth and fusion occurring earlier than palatogenesis. Supporting this distinction, the H131Q variant has been shown to alter *MAFB*-dependent regulation of *ARHGAP29* in a cell-type specific manner, significantly increasing ARHGAP29 in ectodermal-derived mouse cells and also eliminating *ARHGAP29* promoter and enhancer-driven luciferase activity in human mesenchymal cells^57^. Thus, regulatory variation at the *MAFB* locus may influence pathways relevant to lip morphogenesis rather than palatogenesis. Notably, the OFC-associated variant experimentally tested was found in a pooled phenotype, nsCL/P, meaning that CL, specifically, could be a driver for this association.

Further, our present study indicates there is an additional component to left-sided CL bias involved with potential regulation of *MAFB*. Our lead SNP is positioned in a cCRE distal enhancer and mapped to eQTL activity reported for Elastin Microfibril Interfacer 3 (*EMILIN3*), a protein-coding gene involved in elastic fiber formation and collagen chain trimerization pathways. Experimental work demonstrated expression of *Emilin*-3 in the mouse skeleton^58,59^ and expression of zebrafish *EMILIN3* orthologs^58^ localized to cartilage precursors in facial skeletal formation. Overall, these observations align with genetic evidence implicating *MAFB* primarily in CL-related risk. Although genetic modifier tests here additionally provide support for left-sided bias, future confirmatory work will be required to boost power for RCL subtypes and study other broad classes of OFC beyond isolated CL only.

OFCs are complex and deeply heterogeneous with subtype traits manifesting within broad categories, including CL. Indeed, it is common and necessary to pool these subtype traits into broad categories to increase sample size and statistical power. However, as our study demonstrates, future work may benefit from more granular characterizations of OFC phenotypes. The genetic findings gained from our subtype specific analyses provide evidence for distinct genetic factors and developmental pathways involved in the development of oral structures. By pinpointing these genetic contributors, our findings may help improve polygenic risk scores for more accurate prediction of CL and its subtypes. They may also lay the groundwork for possible new therapeutic strategies for cleft repair. This may be particularly relevant for alveolar clefts, which require staged surgeries and autologous bone grafting through early adolescence. Similar to the use of recombinant bone morphogenetic proteins (BMP) instead of grafting in patients with difficult-to-repair alveolar clefts^60,61^, our findings may provide potential molecular targets for inducing alveolar fusion. While functional follow-up is essential to illuminate their biological function, these insights and similar follow-up work may ultimately offer broader understanding of relevance to regenerative medicine and wound healing.

## DECLARATIONS

## Conflict of Interest

The authors have no conflict of interest to declare.

## Data Availability Statement

The GMKF data used in this study is provided in Table S1, along with links to the specific GMKF project and dbGaP accession numbers if the data has been released. There is one GMKF project still under an embargo on dbGaP as of this writing.

## Ethical Oversight

All analyses in this study used publicly available or soon-to-be publicly available data. Written informed consent or assent was provided by all participants prior to participation in any research activities. Details on ethic committees, IRB and FWA numbers are as follows for the following international recruitment sites─ USA: The University of Pittsburgh Institutional Review Board, FWA00006790, Coordinating Center and PA sites approval; The University of Iowa Institutional Review Board, FWA00003007, Coordinating Center and IA project sites approval; University of Texas Houston Health Sciences IRB, FWA00000667, TX project approval; University of Puerto Rico, Medical Sciences Ethical Committee, FWA0000556, PR project approval; Emory University, Biomedical IRB, FWA00005792, GA project approval; Children’s Hospital of Philadelphia (CHOP), FWA00000459, CHOP project approval; ─COLOMBIA: Ethics Committee of Fundacion Clinica Noel de Medellin (no FWA acquired but not required for foreign recruitment sites); ─ARGENTINA: ECLAMC/CEMIC Ethics Committees, Buenos Aires, Argentina, FWA0001566 and FWA00001745; ─GUATEMALA: Ethics Committees of the Hospital Nacional de Huehuetenango and Hospital Nacional Tiquisate, Guatemala (no FWA acquired but not required for foreign recruitment sites); HUNGARY: GAT Foundation Budapest, Hungary, FWA0001328; ─NIGERIA: Lagos University Teaching Hospital, Health Research Ethics Committee, Lagos, Nigeria (no FWA acquired but not required for foreign recruitment sites); ─The PHILIPPINES: University of the Philippines, Manila, Ethics Committee, FWA00003505.

## Funding

This work was funded by the following grants from the National Institute of Health: T90-DE030853 (NH, ZE-Y); X01-HL132363 (MLM, EF); X01-HL136465 (MLM, EF); X01-HD100701 (EJL-C, MLM, JCM); X01-DE030062 (MLM, EF); X01-DE032472 (MLM, EJL-C); T32-GM149422 (GB); R01-DE032122 (JRS); R01-DE032319 (MLM); R01-DE016148 (MLM, SMW); R01-DE031855 (IR, MLM); R01-DE008559 (JCM, MLM); R01-DE028342 (EJL-C). This research was supported in part by the University of Pittsburgh Center for Research Computing and Data, RRID:SCR_022735, through the resources provided. Specifically, this work used the HTC cluster, which is supported by NIH award number S10OD028483.

## CREDiT

Conceptualization: MLM, JRS, SMW, EJL-C, NH; Data curation: NH, ZE-Y, MKL, NM, SWC, SB, GB, TB, AES, JA; Formal analysis: NH, ZE-Y, MKL; Funding acquisition: MLM, EF, EJL-C, IR, JCM, JRS, SMW; Investigation: NH, ZE-Y, MKL, MLM, JRS; Methodology: NH, ZE-Y, MKL; Resources: MLM, JRS, SMW, EJL-C, IR, JCC, JCM, GW, CDP, LMMU, EL, JTH, CJB-M, AB, TB; Software:; Supervision: MLM, JRS; Validation: NH, ZE-Y, MKL; Visualization: NH, ZE-Y; Writing – original draft: NH, ZE-Y; Writing – review & editing: NH, ZE-Y, MKL, MLM, JRS, SMW, EJL-C, IR, SCW, SB, JCC.

## Notes

### Competing Interest Statement

The authors have declared no competing interest.

### Author Declarations

All analyses in this study used publicly available or soon-to-be publicly available data from the Gabriella Miller Kids First Pediatric Research Program. Written informed consent or assent was provided by all participants prior to participation in any research activities. Details on ethic committees, IRB and FWA numbers are as follows for the following international recruitment sites─ USA: The University of Pittsburgh Institutional Review Board, FWA00006790, Coordinating Center and PA sites approval; The University of Iowa Institutional Review Board, FWA00003007, Coordinating Center and IA project sites approval; University of Texas Houston Health Sciences IRB, FWA00000667, TX project approval; University of Puerto Rico, Medical Sciences Ethical Committee, FWA0000556, PR project approval; Emory University, Biomedical IRB, FWA00005792, GA project approval; Children's Hospital of Philadelphia (CHOP), FWA00000459, CHOP project approval; ─COLOMBIA: Ethics Committee of Fundacion Clinica Noel de Medellin (no FWA acquired but not required for foreign recruitment sites); ─ARGENTINA: ECLAMC/CEMIC Ethics Committees, Buenos Aires, Argentina, FWA0001566 and FWA00001745; ─GUATEMALA: Ethics Committees of the Hospital Nacional de Huehuetenango and Hospital Nacional Tiquisate, Guatemala (no FWA acquired but not required for foreign recruitment sites); HUNGARY: GAT Foundation Budapest, Hungary, FWA0001328; ─NIGERIA: Lagos University Teaching Hospital, Health Research Ethics Committee, Lagos, Nigeria (no FWA acquired but not required for foreign recruitment sites); ─The PHILIPPINES: University of the Philippines, Manila, Ethics Committee, FWA00003505.

## REFERENCES

1 Dixon, M. J., Marazita, M. L., Beaty, T. H. & Murray, J. C. Cleft lip and palate: understanding genetic and environmental influences. Nature Reviews Genetics 12, 167–178 (2011).

2 Murray, J. Gene/environment causes of cleft lip and/or palate. Clinical genetics 61, 248–256 (2002).

3 Rahimov, F. et al. Disruption of an AP-2α binding site in an IRF6 enhancer is associated with cleft lip. Nature genetics 40, 1341–1347 (2008).

4 Mitchell, L. E. Twin studies in oral cleft research. Cleft Lip and palate: from origin to treatment 214, 221 (2002).

5 Sivertsen, Å. et al. Familial risk of oral clefts by morphological type and severity: population based cohort study of first degree relatives. Bmj 336, 432–434 (2008).

6 Grosen, D. et al. A cohort study of recurrence patterns among more than 54 000 relatives of oral cleft cases in Denmark: support for the multifactorial threshold model of inheritance. Journal of medical genetics 47, 162–168 (2010).

7 Birnbaum, S. et al. Key susceptibility locus for nonsyndromic cleft lip with or without cleft palate on chromosome 8q24. Nature genetics 41, 473–477 (2009).

8 Mangold, E. et al. Genome-wide association study identifies two susceptibility loci for nonsyndromic cleft lip with or without cleft palate. Nature genetics 42, 24–26 (2010).

9 Beaty, T. H. et al. A genome-wide association study of cleft lip with and without cleft palate identifies risk variants near MAFB and ABCA4. Nature genetics 42, 525–529 (2010).

10 Ludwig, K. U. et al. Genome-wide meta-analyses of nonsyndromic cleft lip with or without cleft palate identify six new risk loci. Nature genetics 44, 968–971 (2012).

11 Yu, Y. et al. Genome-wide analyses of non-syndromic cleft lip with palate identify 14 novel loci and genetic heterogeneity. Nature communications 8, 14364 (2017).

12 Leslie, E. J. et al. Genome-wide meta-analyses of nonsyndromic orofacial clefts identify novel associations between FOXE1 and all orofacial clefts, and TP63 and cleft lip with or without cleft palate. Human genetics 136, 275–286 (2017).

13 Butali, A. et al. Genomic analyses in African populations identify novel risk loci for cleft palate. Human molecular genetics 28, 1038–1051 (2019).

14 Welzenbach, J. et al. Integrative approaches generate insights into the architecture of non-syndromic cleft lip with or without cleft palate. Human Genetics and Genomics Advances 2 (2021).

15 Mukhopadhyay, N. et al. Genome-wide association study of non-syndromic orofacial clefts in a multiethnic sample of families and controls identifies novel regions. Frontiers in cell and developmental biology 9, 621482 (2021).

16 Jia, Z. et al. Multi-ancestry Genome Wide Association Study Meta-analysis of Non-syndromic Orofacial Clefts. medRxiv, 2024.2012. 2006.24318522 (2024).

17 Dack, K. et al. Genetic heterogeneity and homogeneity among orofacial cleft subtypes: genome-wide association studies in the cleft collective. Human Molecular Genetics 34, 1934–1950 (2025).

18 Carlson, J. C. et al. Variants in CALD1, ESRP1, and RBFOX1 are associated with orofacial cleft risk. PLoS genetics 21, e1011581 (2025).

19 Marazita, M. L. The evolution of human genetic studies of cleft lip and cleft palate. Annual review of genomics and human genetics 13, 263–283 (2012).

20 Curtis, S. W. et al. FAT4 identified as a potential modifier of orofacial cleft laterality. Genetic epidemiology 45, 721–735 (2021).

21 Mossey, P. A., Little, J., Munger, R. G., Dixon, M. J. & Shaw, W. C. Cleft lip and palate. The Lancet 374, 1773–1785 (2009).

22 Nasreddine, G., El Hajj, J. & Ghassibe-Sabbagh, M. Orofacial clefts embryology, classification, epidemiology, and genetics. Mutation Research/Reviews in Mutation Research 787, 108373 (2021).

23 Leslie, E. J. et al. A multi-ethnic genome-wide association study identifies novel loci for non-syndromic cleft lip with or without cleft palate on 2p24. 2, 17q23 and 19q13. Human molecular genetics 25, 2862–2872 (2016).

24 Alade, A. et al. Shared genetic risk between major orofacial cleft phenotypes in an African population. Genetic epidemiology 48, 258–269 (2024).

25 Robinson, K. et al. Trio-based GWAS identifies novel associations and subtype-specific risk factors for cleft palate. Human Genetics and Genomics Advances 4 (2023).

26 Gowans, L. J. et al. Genome-wide scan for parent-of-origin effects in a sub-Saharan African cohort with nonsyndromic cleft lip and/or cleft palate (CL/P). The Cleft Palate Craniofacial Journal 59, 841–851 (2022).

27 Huang, L. et al. Genetic factors define CPO and CLO subtypes of nonsyndromicorofacial cleft. PLoS genetics 15, e1008357 (2019).

28 Carlson, J. C. et al. Identification of 16q21 as a modifier of nonsyndromic orofacial cleft phenotypes. Genetic epidemiology 41, 887–897 (2017).

29 Robinson, K., Curtis, S. W. & Leslie, E. J. The heterogeneous genetic architectures of orofacial clefts. Trends in Genetics 40, 410–421 (2024).

30 Curtis, S. W. et al. The PAX1 locus at 20p11 is a potential genetic modifier for bilateral cleft lip. Human Genetics and Genomics Advances 2 (2021).

31 Carlson, J. C. et al. A systematic genetic analysis and visualization of phenotypic heterogeneity among orofacial cleft GWAS signals. Genetic epidemiology 43, 704–716 (2019).

32 Ray, D. et al. Pleiotropy method reveals genetic overlap between orofacial clefts at multiple novel loci from GWAS of multi-ethnic trios. PLoS genetics 17, e1009584 (2021).

33 Mukhopadhyay, N. et al. Genome-wide association study of multiethnic nonsyndromic orofacial cleft families identifies novel loci specific to family and phenotypic subtypes. Genetic epidemiology 46, 182–198 (2022).

34 Duan, S., Shi, J., Shi, B. & Jia, Z. Association analysis of GWAS hits and non-syndromic cleft lip with/without palate with cleft alveolar in Han population of western China. International Journal of Clinical and Experimental Pathology 13, 2576 (2020).

35 Wurfbain, L. F. et al. Diagnostic gene panel testing in (non)-syndromic patients with cleft lip, alveolus and/or palate in the Netherlands. Molecular Syndromology 14, 270–282 (2023).

36 Carlson, J. C. et al. Identifying genetic sources of phenotypic heterogeneity in orofacial clefts by targeted sequencing. Birth defects research 109, 1030–1038 (2017).

37 Kriens, O. LAHSHAL: a concise documentation system for cleft lip, alveolus, and palate diagnoses. What is a cleft lip and palate, 32–33 (1989).

38 Houkes, R. P. et al. Learnability of the LAHSHAL Classification for Oral Clefts: Results of an International Webinar. Journal of Craniofacial Surgery, 10.1097 (2025).

39 DePristo, M. A. et al. A framework for variation discovery and genotyping using next-generation DNA sequencing data. Nature genetics 43, 491–498 (2011).

40 McKenna, A. et al. The Genome Analysis Toolkit: a MapReduce framework for analyzing next-generation DNA sequencing data. Genome research 20, 1297–1303 (2010).

41 Van der Auwera, G. A. et al. From FastQ data to high-confidence variant calls: the genome analysis toolkit best practices pipeline. Current protocols in bioinformatics 43, 11.10.11–11.10.33 (2013).

42 Chang, C. C. et al. Second-generation PLINK: rising to the challenge of larger and richer datasets. GigaScience 4 (2015). 10.1186/s13742-015-0047-8

43 Manichaikul, A. et al. Robust relationship inference in genome-wide association studies. Bioinformatics 26, 2867–2873 (2010).

44 He, Y., Koido, M., Shimmori, Y. & Kamatani, Y. GWASLab: a Python package for processing and visualizing GWAS summary statistics. (2023).

45 Watanabe, K., Taskesen, E., Van Bochoven, A. & Posthuma, D. Functional mapping and annotation of genetic associations with FUMA. Nature communications 8, 1826 (2017).

46 Wang, G., Sarkar, A., Carbonetto, P. & Stephens, M. A simple new approach to variable selection in regression, with application to genetic fine mapping. Journal of the Royal Statistical Society Series B: Statistical Methodology 82, 1273–1300 (2020).

47 The 1000 Genomes Project Consortium. A global reference for human genetic variation. Nature 526, 68–74 (2015). 10.1038/nature15393

48 Kanai, S. M. et al. Auriculocondylar syndrome 2 results from the dominant-negative action of PLCB4 variants. Disease Models & Mechanisms 15, dmm049320 (2022).

49 Rieder, M. J. et al. A human homeotic transformation resulting from mutations in PLCB4 and GNAI3 causes auriculocondylar syndrome. The American Journal of Human Genetics 90, 907–914 (2012).

50 Daisy, C. S. et al. Expression and localization of Phosphoinositide-specific Phospholipases C in cultured, differentiating and stimulated human osteoblasts. Journal of Cellular Signaling 3, 44–61 (2022).

51 Jakovljevic, A. et al. Involvement of the Notch signaling system in alveolar bone resorption. Japanese Dental Science Review 59, 38–47 (2023).

52 Omi, M. & Mishina, Y. Roles of osteoclasts in alveolar bone remodeling. genesis 60, e23490 (2022).

53 Smith, W., Markus, A. & Delaire, J. Primary closure of the cleft alveolus: a functional approach. British Journal of Oral and Maxillofacial Surgery 33, 156–165 (1995).

54 Lennon, C. J. et al. Association of candidate genes with nonsyndromic clefts in Honduran and Colombian populations. The Laryngoscope 122, 2082–2087 (2012).

55 Pan, Y. et al. Different roles of two novel susceptibility loci for nonsyndromic orofacial clefts in a Chinese Han population. American Journal of Medical Genetics Part A 155, 2180–2185 (2011).

56 Leslie, E. J. & Marazita, M. L. in American Journal of Medical Genetics Part C: Seminars in Medical Genetics. 246–258 (Wiley Online Library).

57 Paul, B. J. et al. The Mafb cleft-associated variant H131Q is not required for palatogenesis in the mouse. Developmental Dynamics 250, 1463–1476 (2021).

58 Schiavinato, A. et al. EMILIN-3, peculiar member of elastin microfibril interface-located protein (EMILIN) family, has distinct expression pattern, forms oligomeric assemblies, and serves as transforming growth factor β (TGF-β) antagonist. Journal of Biological Chemistry 287, 11498–11515 (2012).

59 Leimeister, C., Steidl, C., Schumacher, N., Erhard, S. & Gessler, M. Developmental expression and biochemical characterization of Emu family members. Developmental biology 249, 204–218 (2002).

60 Clark, M. & Cunningham, L. Effect of Recombinant Human Bone Morphogenetic Protein on Alveolar Grafting of Cleft Lip and Palate Patients. Journal of Oral and Maxillofacial Surgery 72, e13–e14 (2014).

61 Makar, K. G., Buchman, S. R. & Vercler, C. J. Bone morphogenetic protein-2 and demineralized bone matrix in difficult bony reconstructions in cleft patients. Plastic and Reconstructive Surgery–Global Open 9, e3611 (2021).

