## Supplementary Figures for "Trio-based GWAS reveals novel loci associated with different forms of isolated cleft lip"

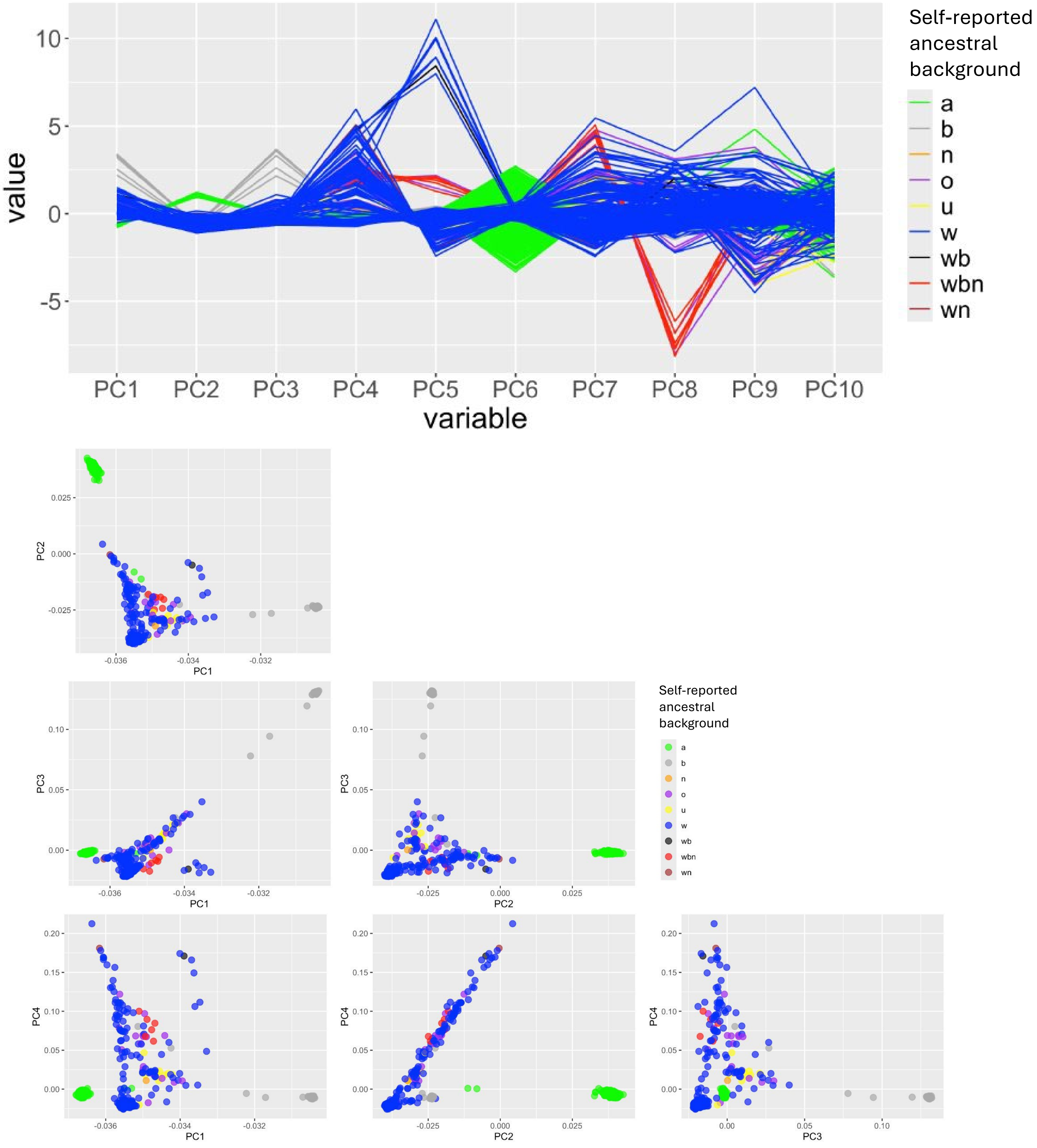


**Figure S1** Genetic PCA plots of all individuals in the study and labeled with self-reported ancestral background nomenclature used in the collection documents.

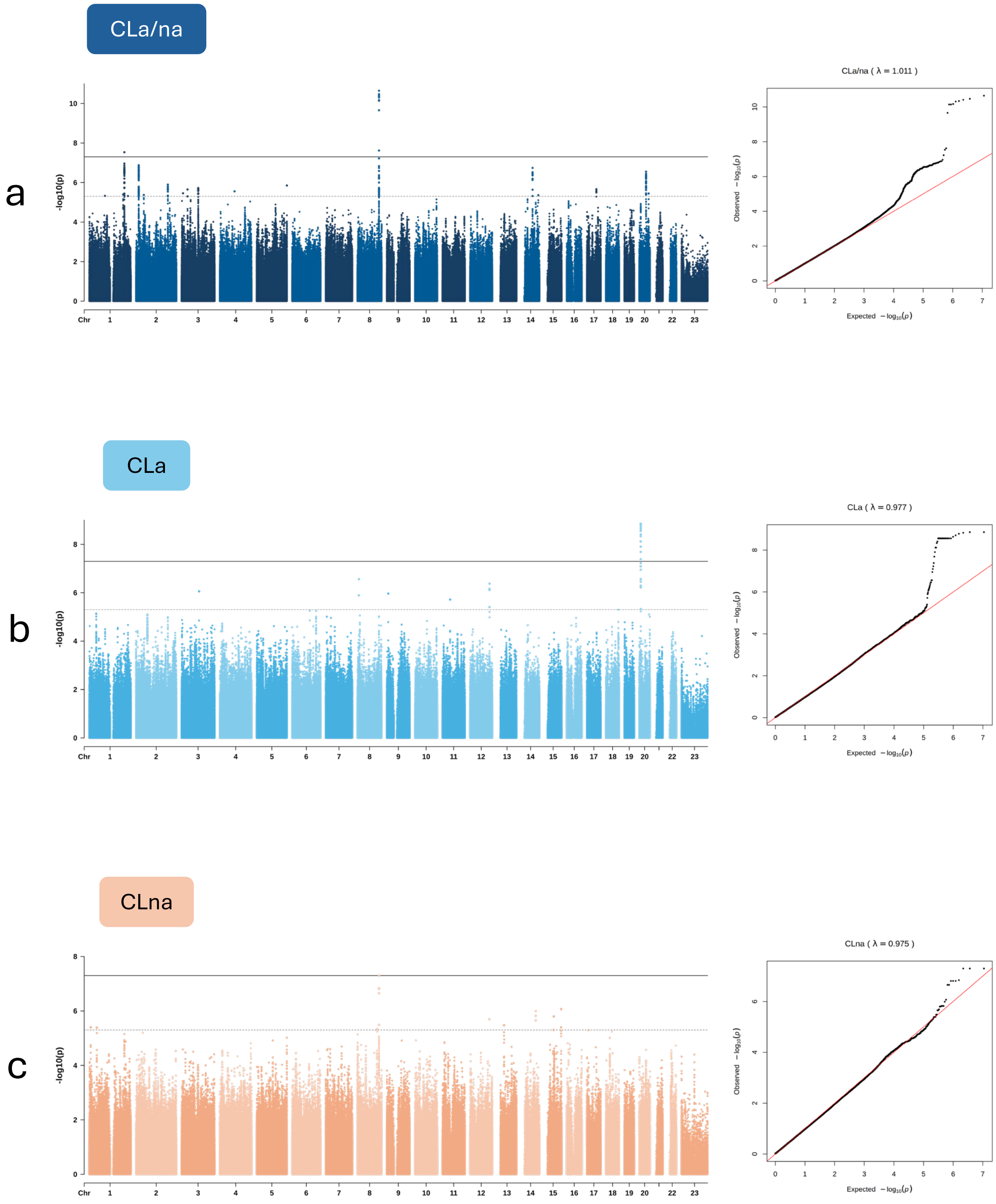


**Figure S2** Manhattan plots and QQ plots of the GWASs for the three primary groupings of CL in this study: **a)** CLa/na, **b)** CLa, and **c)** CLna.

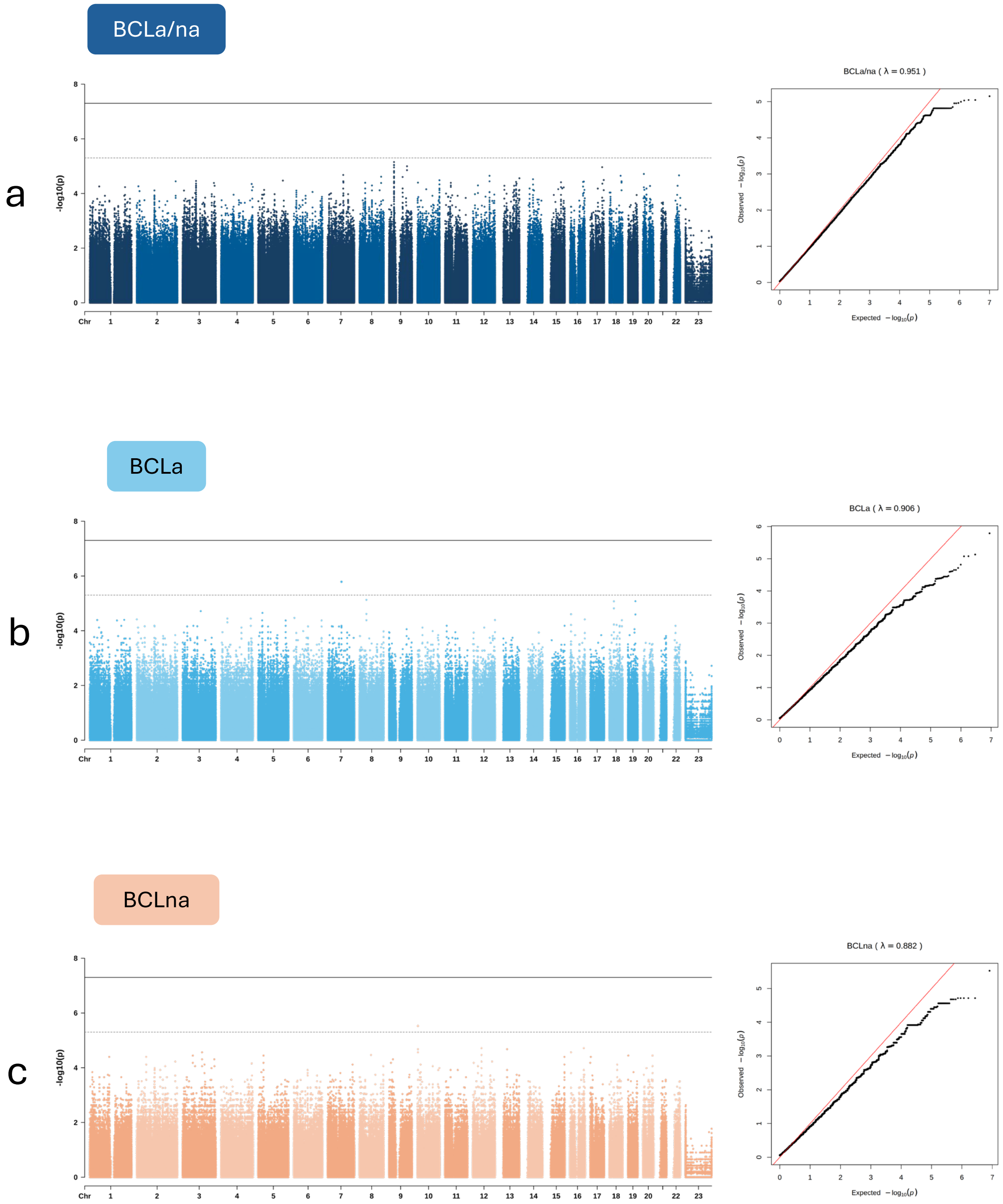


**Figure S3** Manhattan plots and QQ plots of the GWASs for the BCL subtype groupings of CL in this study: **a)** BCLa/na, **b)** BCLa, and **c)** BCLna.

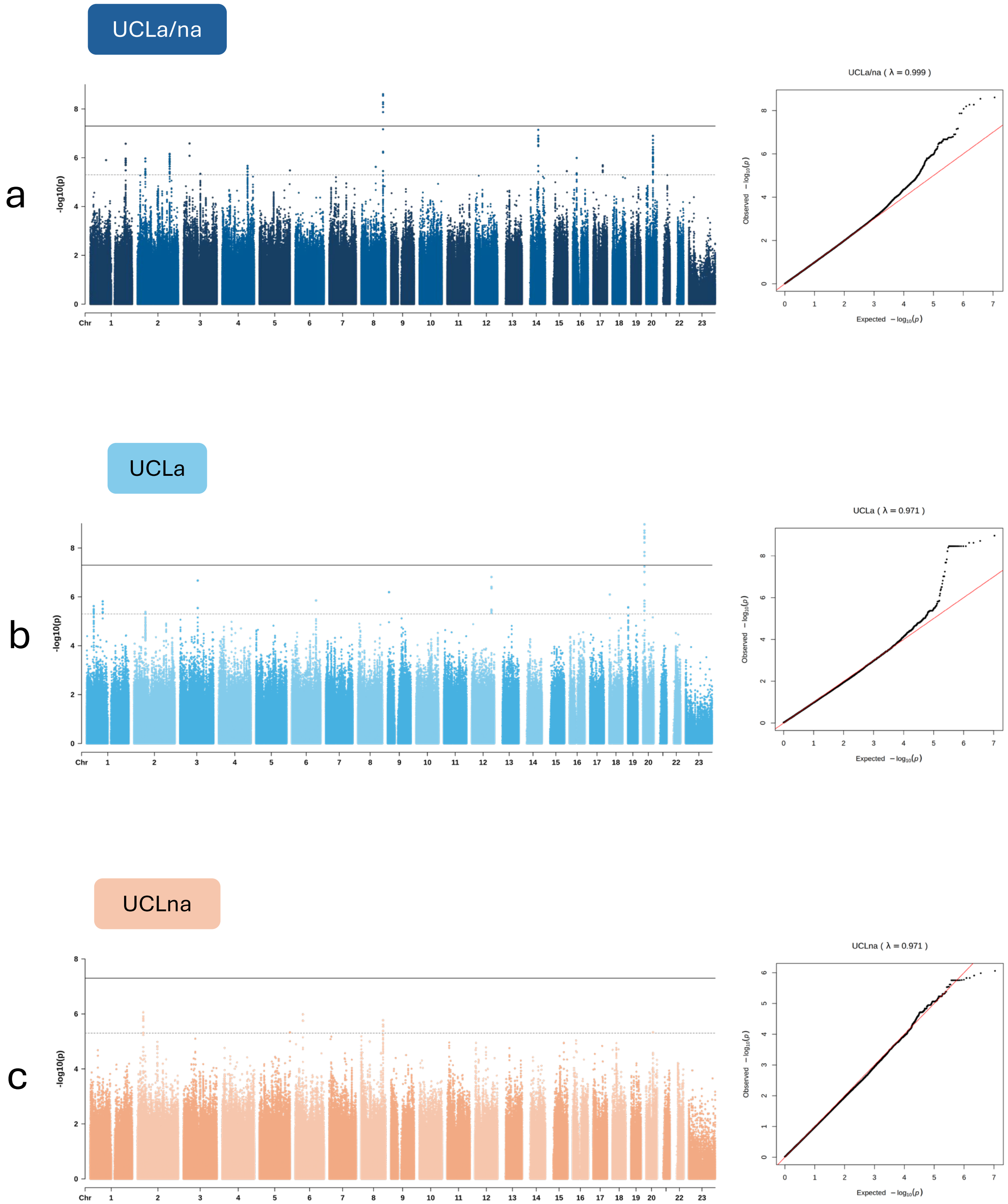


**Figure S4** Manhattan plots and QQ plots of the GWASs for the UCL subtype groupings of CL in this study: **a)** UCLa/na, **b)** UCLa, and **c)** UCLna.

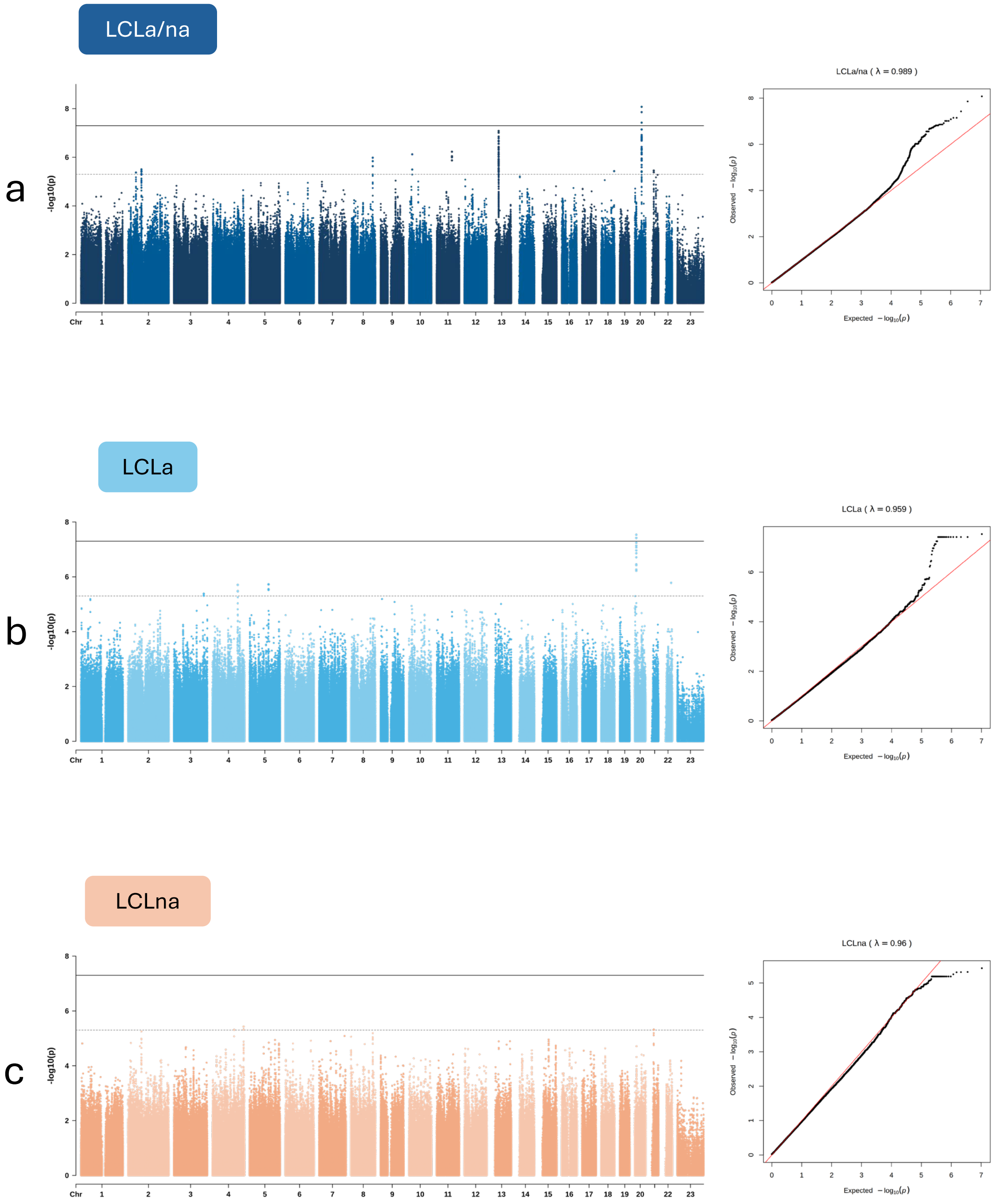


**Figure S5** Manhattan plots and QQ plots of the GWASs for the LCL subtype groupings of CL in this study: **a)** LCLa/na, **b)** LCLa, and **c)** LCLna.

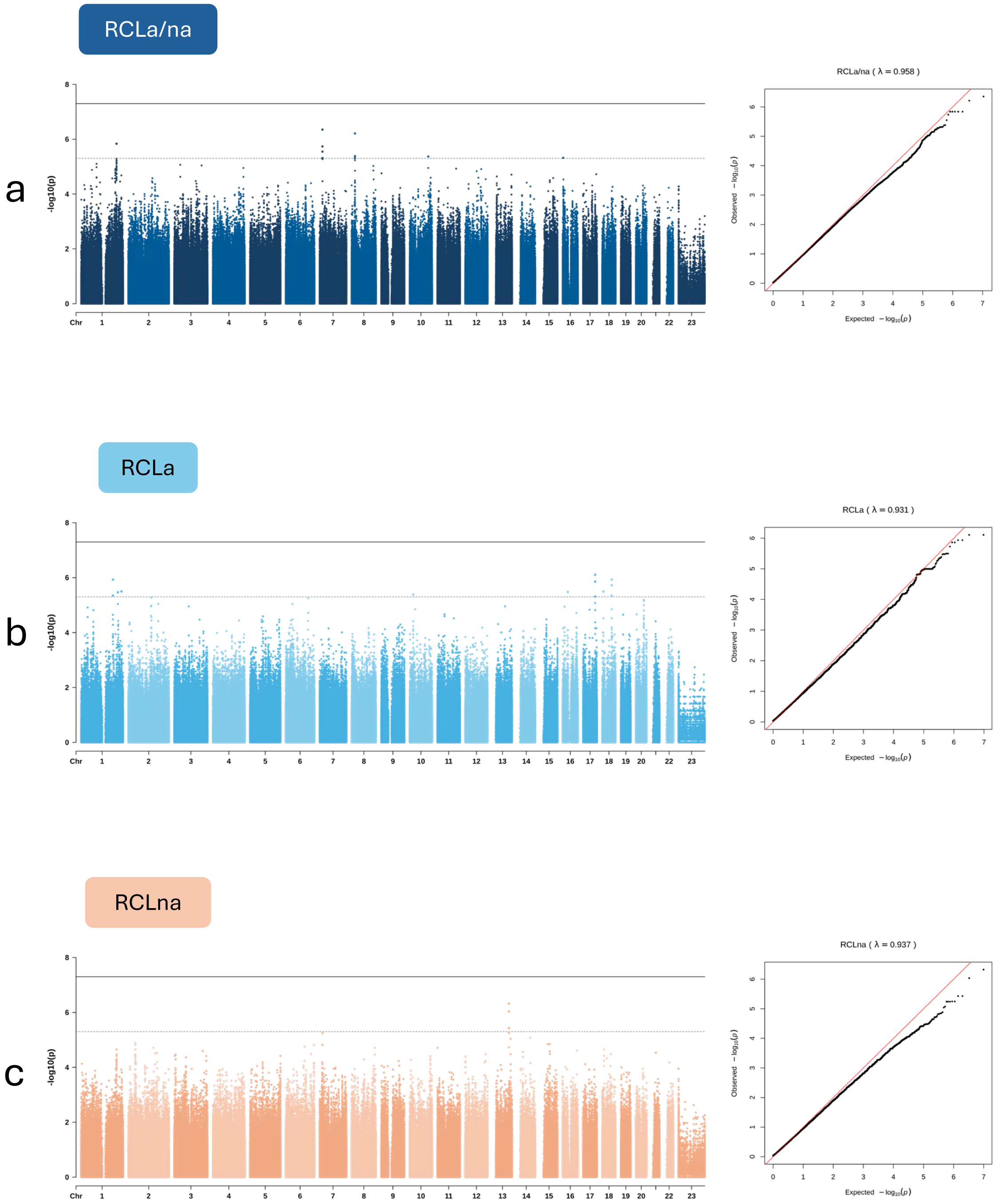


**Figure S6** Manhattan plots and QQ plots of the GWASs for the RCL subtype groupings of CL in this study: **a)** RCLa/na, **b)** RCLa, and **c)** RCLna.

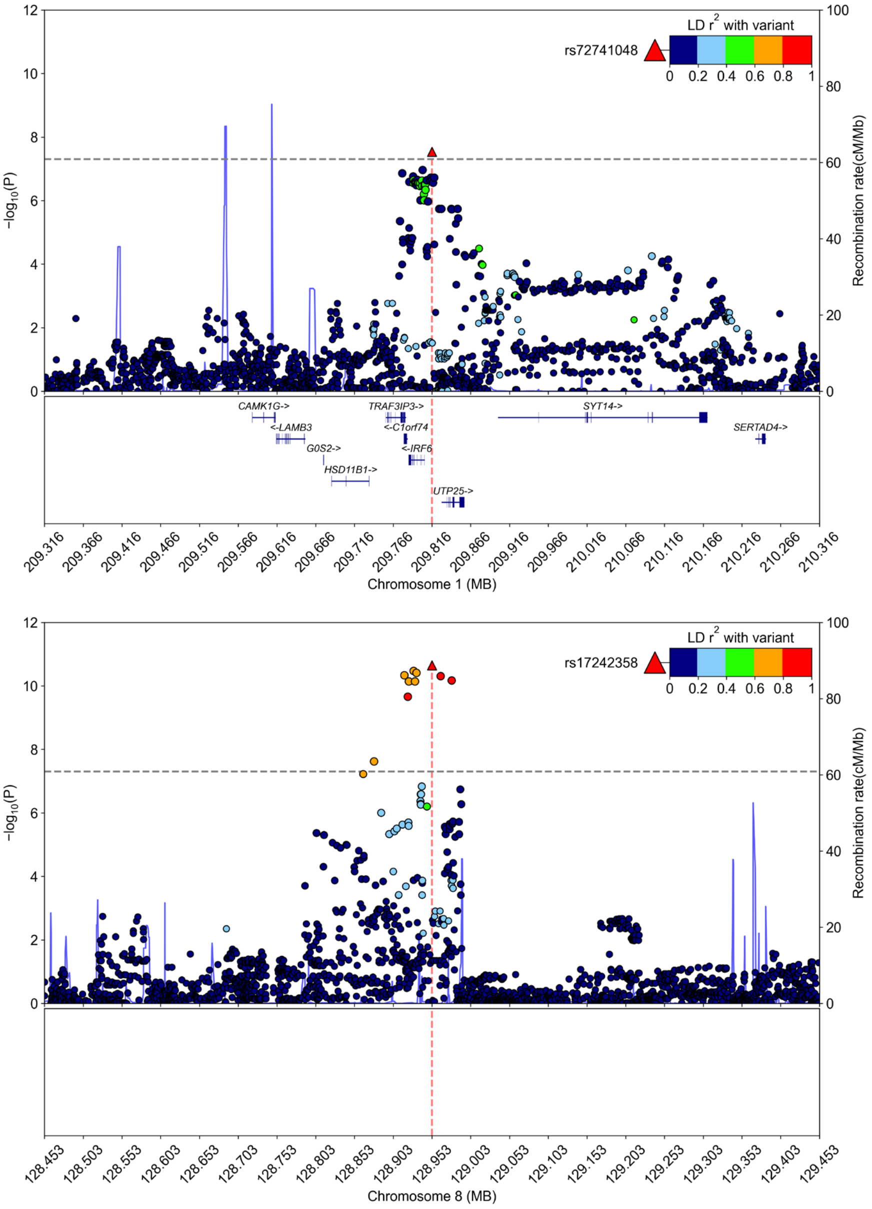


**Figure S7** Regional association plots of significant loci **a)** *IRF6* and **b)** 8q24.21 from the CLa/na (n=837) trio-based GWAS.

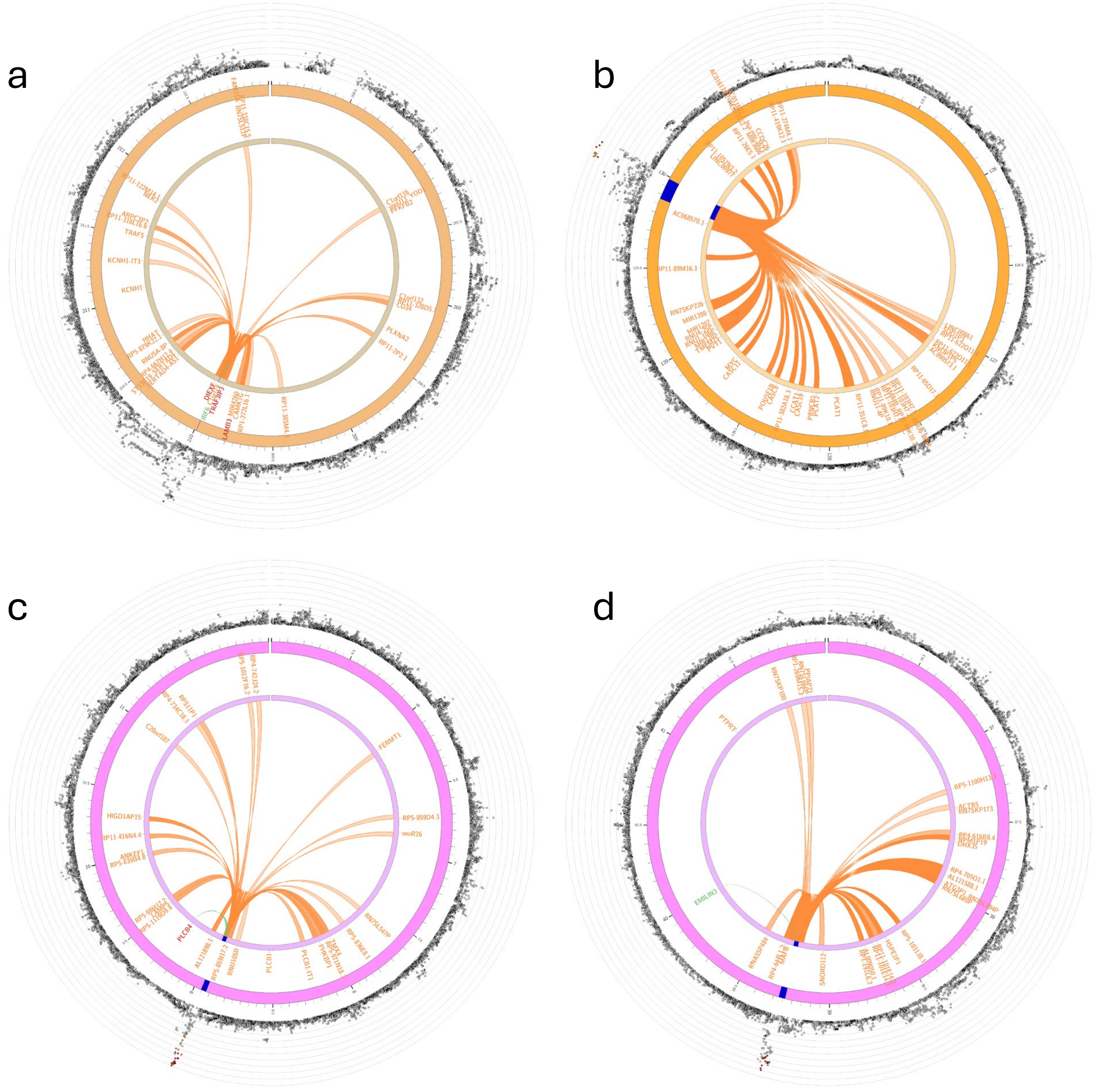


**Figure S8** FUMA-based gene mapping plots for the four lead SNPs in our study. **a)** rs72741048, **b)** rs17242358, **c)** rs2206265, **d)** rs6065259. The strings and color of string extending from the SNP position represent mapping strategies: eQTL data (green), chromatin interaction (orange), and both (red). The third mapping strategy was based on genomic position and is indicated by the nearness of the SNP. The SNPs (points) extending on the outer rings are colored by linkage disequilibrium.

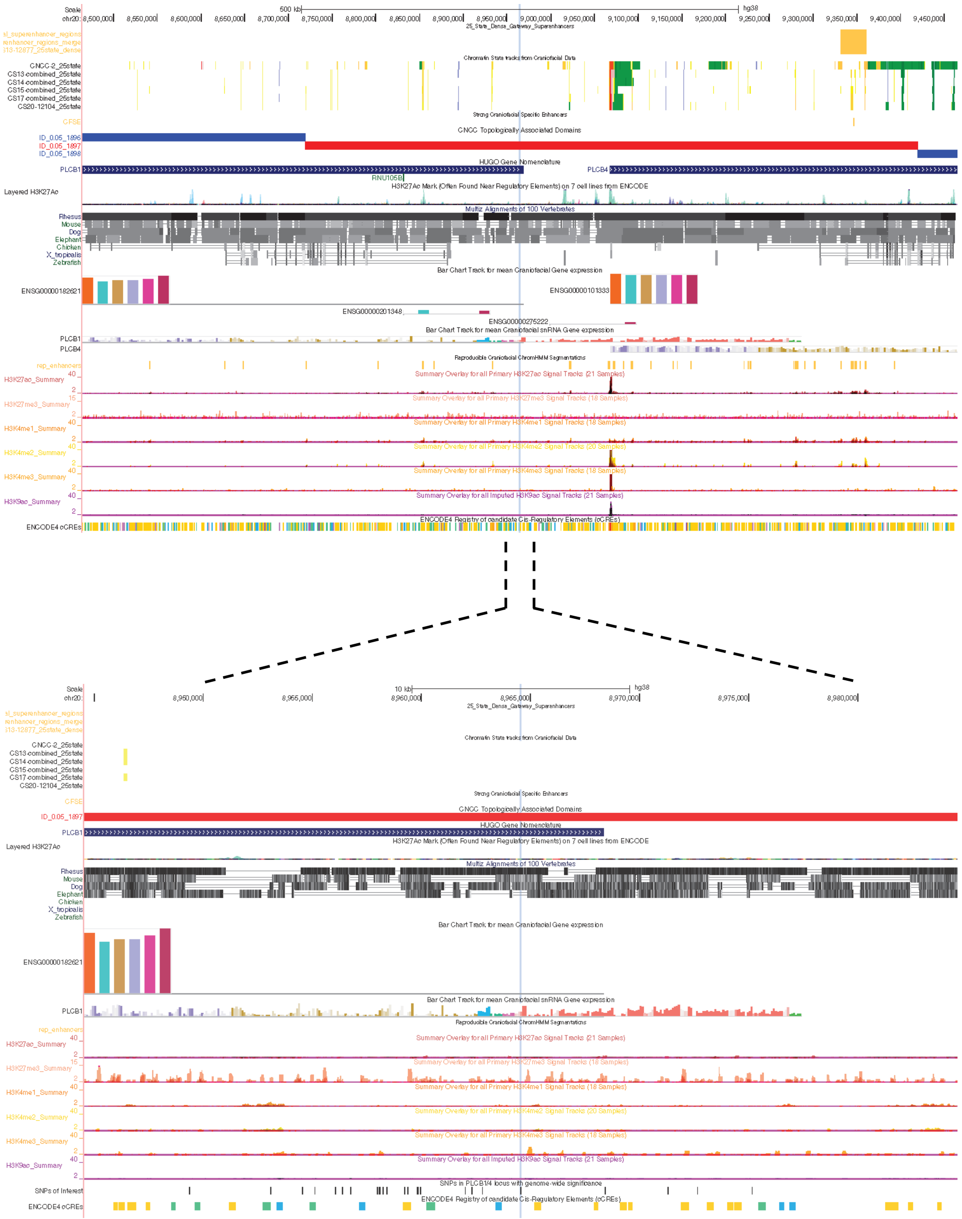


**Figure S9** UCSC Genome browser view of the *PLCB1*/*PLCB4* locus. The lead SNP, rs2206265, is indicated by the vertical blue highlighted line.

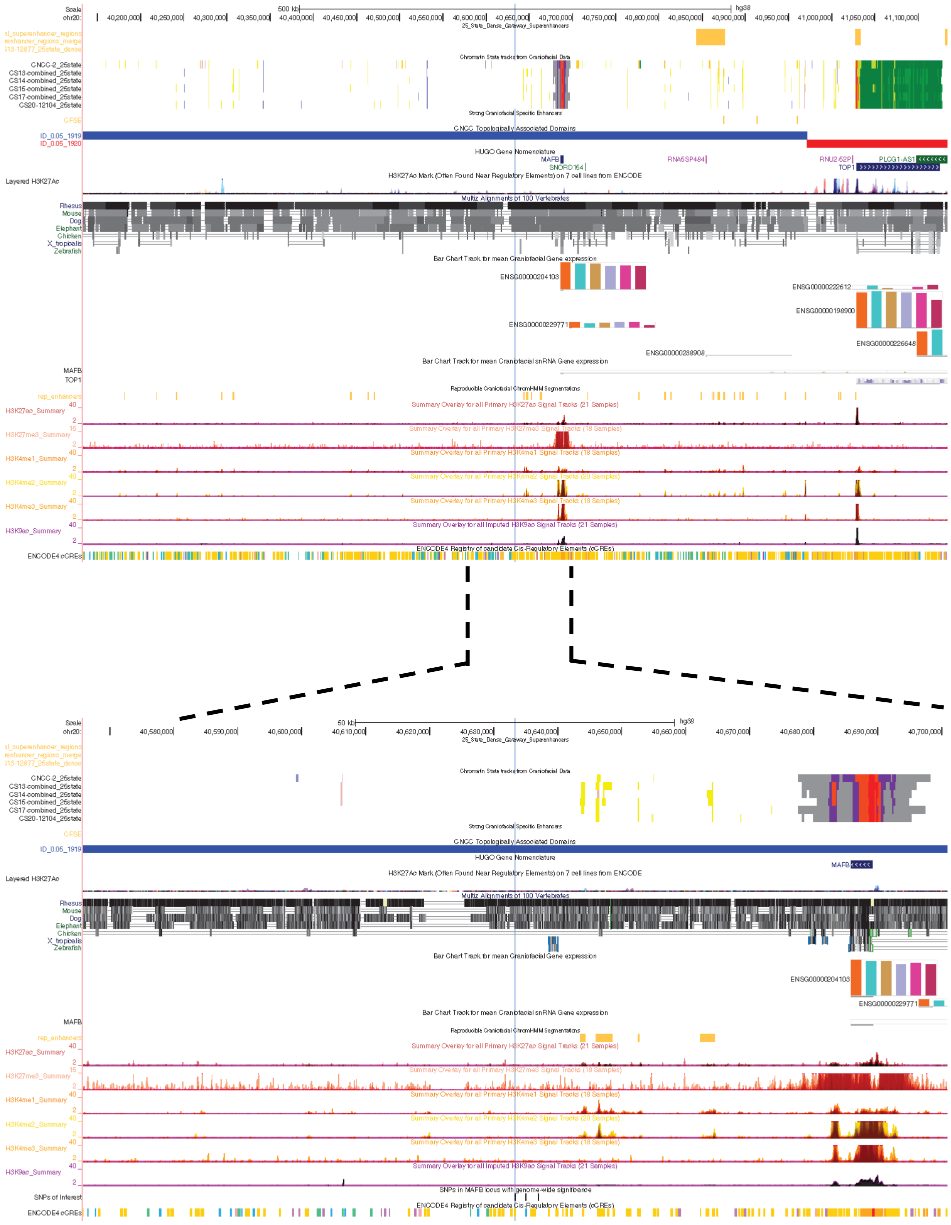

**Figure S10** UCSC Genome browser view of the *MAFB* locus. The lead SNP, rs6065259, is indicated by the vertical blue highlighted line.
